# Dementia Risk Following Ischemic Stroke: A Systematic Review and Meta-Analysis of Factors Collected at Time of Stroke Diagnosis

**DOI:** 10.1101/2021.10.28.21265621

**Authors:** Reem Waziry, Jacqueline J Claus, Albert Hofman

## Abstract

**Objective:** To assess incidence rates and predictors of dementia after ischemic stroke.

**Methods:** A search was conducted on Embase and Medline for reports published up to November 2019. Studies were included if they: 1) assessed dementia incidence among patients with ischemic stroke diagnosis and 2) excluded patients with prevalent dementia at baseline. The main analysis included: 1) absolute risk; 2) incidence rates (per 100 person-years) and 3) patient-level predictors (demographics, CVD history and major cardiac events, previous stroke and TIA, stroke location, disability post-stroke, chronic brain change and stroke mechanism). Additional predictors assessed included study setting (clinic or registry), method of dementia diagnosis (Diagnostic and Statistical Manual of Mental Disorders (DSM), National Institute of Neurological Disorders and Stroke (NINDS) or both) and inclusion of patients with recurrent or first-ever stroke. A random effects meta-analysis was undertaken. Risk of bias in included studies was assessed in terms of selection, comparability and outcome.

**Results:** 4,325 studies were screened in the title and abstract phase after removing duplicates and 280 eligible studies were screened for full text. A total of 21 studies met the inclusion criteria and were included in the meta-analysis, representing 55,183 patients with ischemic stroke, with average age of 70 years (range 65-80 years) and average follow-up of 29 months. The majority of included studies were conducted in a hospital setting (n=17/21). The overall rate of dementia after ischemic stroke was 13.0 per 1000 person-years (95% CI 6.0, 36.0). Incidence rates were eight times higher in hospital-based studies (17.0, 95% CI 8.0, 36.0) compared to registry-based studies (1.8, 95% CI 0.8, 4.0). Absolute dementia risk after stroke was 20% at 5 year, 30% at 15 years and 48% at 25 years of follow-up. Incidence rates were 1.5 times higher in studies that included patients with recurrent ischemic stroke compared to estimates from studies that included first-time ever stroke patients only. There was 33% difference in dementia incidence in the later study periods (2007 – 2009) compared to (1996 – 2006). Statistically significant predictors of dementia after ischemic stroke included female gender (OR=1.2, 95% CI 1.1, 1.4), hypertension (1.4, 95% 1.1, 2.0), diabetes mellitus (1.6, 95% 1.3, 2.1), atrial fibrillation (1.9, 95% 1.2, 3.0), previous stroke (2.0, 95% CI 1.6, 2.6), presence of stroke lesion in dominant hemisphere (2.4, 95% 1.3, 4.5), brain stem/cerebellum (0.5, 95% CI 0.3, 0.9) or frontal lobe (3.7, 95% CI 1.2, 12.0), presence of aphasia (7.9, 95% CI 2.4, 26.0), dysphasia (5.8, 95% CI 3.0, 11.3), gait impairment (1.7, 95% CI 1.1, 2.7), presence of white matter hyperintensities (3.2, 95% CI 2.0, 5.3), medial temporal lobe atrophy (3.9, 95% CI 1.9, 8.3) and transient ischemic attack (TIA) as the predisposing aetiology for ischemic stroke (0.44, 95% CI 0.22, 0.88).

**Conclusion:** Factors routinely collected at time of admission guide informed monitoring of patients at highest risk of progression to dementia after acute ischemic stroke. Predictors of dementia after acute ischemic stroke should be assessed as distinct features from those established for general dementia.

**Key Points:** *Question:* What is the likelihood of progression to dementia after ischemic stroke? And what are the individual predictors?

*Findings:* The overall rate of dementia after ischemic stroke was 13.0 per 1000 person-years (95% CI 6.0, 36.0). Incidence rates were eight times higher in hospital-based studies (17.0, 95% CI 8.0, 36.0) compared to registry-based studies (1.8, 95% CI 0.8, 4.0). Absolute dementia risk after stroke was 20% at 5 years, 30% at 15 years and 48% at 25 years of follow-up. Incidence rates were 1.5 times higher in studies that included patients with recurrent ischemic stroke compared to estimates from studies that included first-time ever stroke patients only. There was 33% difference in dementia incidence in the later study periods (2007 – 2009) compared to (1996 – 2006). Statistically significant predictors of dementia after ischemic stroke included female gender, hypertension, diabetes mellitus, atrial fibrillation, previous stroke, presence of stroke lesion in dominant hemisphere, brain stem/cerebellum or frontal lobe, presence of aphasia, dysphasia, gait impairment, presence of white matter hyperintensities, medial temporal lobe atrophy and transient ischemic attack (TIA) as the predisposing aetiology for ischemic stroke.

*Meaning:* Factors routinely collected at time of admission guide informed monitoring of patients at highest risk of progression to dementia after acute ischemic stroke. Predictors of dementia after acute ischemic stroke should be assessed as distinct features from those established for general dementia.

## INTRODUCTION

On average, every 40 seconds someone in the United States suffers a stroke ^1^. Stroke survivors represent a high-risk population for progression to cognitive impairment with one in three patients developing dementia after any stroke ^2–5^.

Over the past decade, there has been a lot of emphasis on risk of dementia after any stroke without subtype specification ^6, 7^. Stroke subtypes vary substantially in severity, management, and prognosis, yet combined evidence in this area remains to be very limited with inconsistency in reported estimates in individual studies.

The majority of stroke cases are ischemic in origin. There have been continuing advances in ischemic stroke treatment through timely treatment and counselling. In the present report, we conducted a systematic review and meta-analysis to assess incidence rates and predictors of dementia occurrence after ischemic stroke. We searched for relevant studies published up to 2019 without geographical or setting restrictions.

## METHODS

### Search strategy and Selection Criteria

A comprehensive literature search was conducted on MEDLINE and EMBASE (as of April 1^st^, 2019) using a previously validated search strategy^6^. Searches were limited to studies with human participants. The reference lists of relevant reports and books of abstracts from major recent international stroke and dementia conferences (European Stroke Conference 2018; Alzheimer Association International Conference 2018; International Stroke Conference 2019) were scrutinized for additional studies. This review was conducted according to the Preferred Reporting Items for Systematic Reviews and Meta-Analyses (PRISMA)^8^ [supplementary table 2].

Inclusion and exclusion criteria were determined a priori. Eligible study designs included retrospective or prospective observational cohort studies, randomized controlled trials and interventional studies. Studies were included if they: 1) assessed dementia incidence among patients with ischemic stroke diagnosis and 2) excluded patients with prevalent dementia at baseline. In the case of multiple eligible studies from the same population, the most complete or the most recent respectively were included in the meta-analysis.

### Data extraction and management

Data extraction was completed using a standardized form designed in advance. Study demographic, clinical and epidemiological variables were extracted. A separate dataset was set-up for each predictor to calculate the measures of association with dementia.

### Risk of bias assessment

Risk of bias in observational studies was assessed using a modified version of the Newcastle-Ottawa scale that assesses study quality on the basis of selection, comparability and outcome and ranked as low, high or unclear bias in a similar manner to the Cochrane Risk of Bias tool ^9, 10^.

### Data synthesis and analysis

Characteristics of individual studies were summarized as averages for continuous variables and proportions for categorical variables. The following analyses were assessed: 1) absolute risk; 2) incidence rates and 3) patient-level predictors (demographics, cardiovascular disease (CVD) history and major cardiac events, previous stroke and transient ischemic attacks (TIA), stroke location, disability post-stroke, chronic brain change and stroke mechanism). Additional predictors assessed included study setting (clinic or registry), method of diagnosis (Diagnostic and Statistical Manual of Mental Disorders (DSM), National Institute of Neurological Disorders and Stroke (NINDS) or both) and inclusion of patients with recurrent or first-ever stroke.

Incidence rates calculation was restricted to studies with more than 6 months of follow-up after stroke to minimize counting dementia cases that were likely present before or at the time of ischemic stroke diagnosis. Odds ratios and standard errors were calculated^11^. The inverse variance weighted random effects model was used to pool the log transformed effect estimates and standard errors from primary studies.

## Sensitivity analysis

Sensitivity analyses were performed to assess the influence of each individual study, omitting the studies that had the largest effect on the overall result one by one, to a minimum of three. The *I*^2^ statistic was used to assess heterogeneity between studies^12^. Egger’s test was used to assess publication bias^13^. Analyses were performed using the Stata v14.0 (StataCorp, College Station, Texas).

## RESULTS

### Characteristics of included studies

A total of 5,656 citations were retrieved from the electronic search. After removing duplicates (n=1,331), 4,325 abstracts of citations were reviewed, and 280 citations were eligible for full text screening. A total of 21 studies matched the eligibility criteria and were included in the meta-analysis^14–26^ [Figure 1].

**Figure 1.**
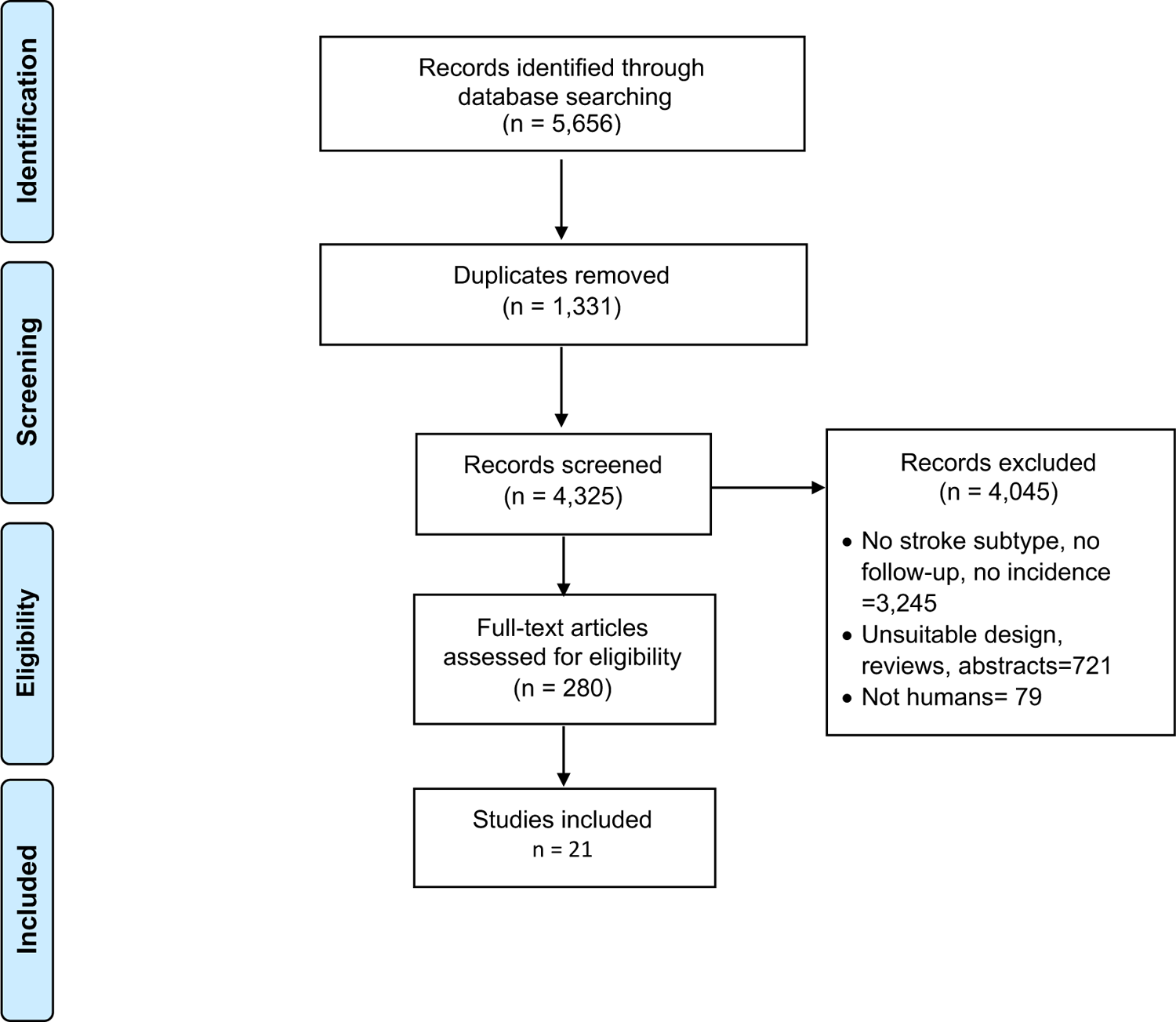
Study flowchart (search updated to November 1^st^, 2019)

The included studies comprised 55,183 patients with ischemic stroke, among which 11,739 dementia cases occurred over an average follow-up of 28.7 months, with 70 years average age at baseline. The majority of included studies were in hospital-based setting (n=17/21), published between 1996 up to 2019 in US, Europe, Asia and Australia. [Figures 1, 2, Table 1].

**Figure 2.**
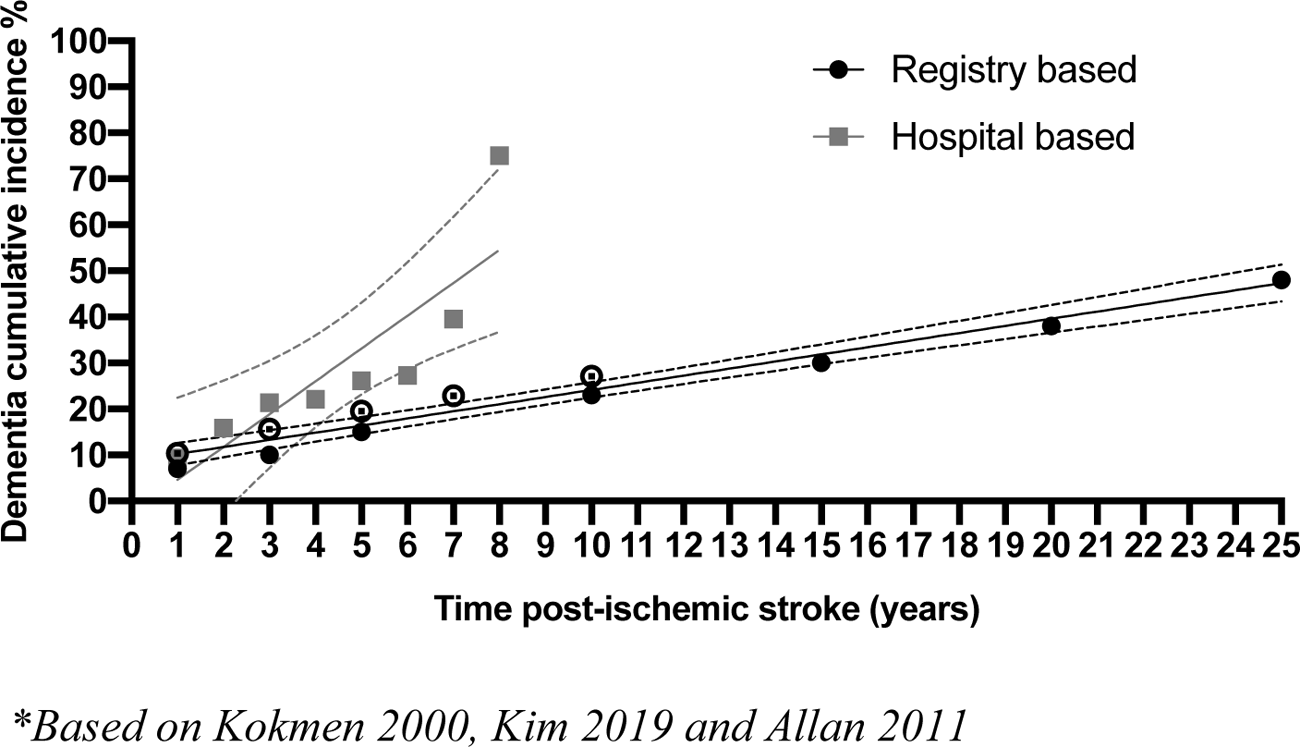
Absolute risk of dementia after ischemic stroke stratified by study setting (hospital vs registry)

**Table 1.** Summary of included studies (N=55,183 ischemic stroke patients)

| Author | Year | Country | Design | Setting | N | Average age | Average follow-up (months) | First-ever stroke | Dementia cases | Dementia diagnosis |
| --- | --- | --- | --- | --- | --- | --- | --- | --- | --- | --- |
| **Censori** | 1996 | Italy | Prospective | Hospital | 104 | 65.0 | 3.0 | Y | 15 | NINDS-AIREN |
| **Kokmen** | 1996 | USA | Retrospective | Registry | 971 | NA | 84.0 | Y | 196 | Medical records |
| **Bornstein** | 1996 | Israel | Prospective | Hospital | 157 | 72.3 | 60.0 | Y | 56 | DSM-III-R |
| **Desmond** | 2000 | USA | Prospective | Hospital | 453 | 70.4 | 21.0 | N | 119 | DSM-III-R |
| **Pohjasvaara** | 2000 | Finland | Prospective | Hospital | 337 | 70.2 | 3.0 | N | 87 | DSM-III |
| **Mok** | 2004 | China | Prospective | Hospital | 75 | 71.0 | 3.0 | N | 10 | CDR |
| **Rasquin** | 2004 | Netherlands | Prospective | Hospital | 176 | 68.0 | 12.0 | Y | 17 | DSM-IV |
| **Zhou** | 2004 | China | Prospective | Hospital | 434 | 69.5 | 3.0 | N | 118 | DSM-IV |
| **Sachdev** | 2006 | Australia | Prospective | Hospital | 104 | 72.2 | 4.0 | N | 36 | By consensus |
| **Gur** | 2010 | Israel, Turkey | Prospective | Hospital | 37 | 76.0 | 4.5 | Y | 17 | DSM-IV/<br>NINDS-AIREN |
| **Allan** | 2011 | UK | Prospective | Hospital | 50 | 80.0 | 45.5 | N | 23 | DSM III-R |
| **Melkas** | 2012 | Finland | Prospective | Hospital | 263 | 70.8 | 90.0 | N | 47 | DSM-IV |
| **Brucki** | 2012 | Brazil | Prospective | Hospital | 172 | 67.7 | 12.0 | N | 21 | NINDS-AIREN |
| **Sibolt** | 2013 | Finland | Prospective | Hospital | 388 | 72.0 | 144.0 | Y | 115 | DSM-III |
| **Tu** | 2014 | China | Cross-sectional | Registry | 689 | 68.6 | 3.0 | N | 67 | NINDS-AIREN |
| **Yang** | 2015 | China | Prospective | Hospital | 1013 | 69.2 | 4.5 | N | 88 | DSM-IV |
| **Mok** | 2016 | China | Prospective | Hospital | 919 | 67.6 | 36.0 | N | 40 | DSM-IV |
| **Makin** | 2018 | UK | Prospective | Registry | 264 | 67.0 | 12.0 | N | 3 | Clinically diagnosed |
| **Surwan** | 2018 | Thailand | Prospective | Hospital | 401 | 63.0 | 6.0 | N | 227 | DSM-V |
| **Kim** | 2019 | Korea | Retrospective | Registry | 47779 | NA | NA | N | 10357 | Medical records |
| **Molad** | 2019 | Israel | Prospective | Hospital | 397 | 69.0 | 24.0 | Y | 80 | MoCA/NeuroTax |

NINDS: National Institute of Neurological Disorders; DSM: Diagnostic and Statistical Manual of Mental Disorders; Association Internationale pour la Recherche et l'Enseignement en Neurosciences; MoCA: Montreal Cognitive Assessment

### Absolute risk of dementia after ischemic stroke

The overall absolute risk of dementia after ischemic stroke was 20% at 5 years, 30% at 15 years and 48% at 25 years of follow-up according to data from three studies. In terms of stratified estimates according to study settings, absolute risk of dementia after ischemic stroke for hospital-based setting was as follows: 1 year (9.4); 3 year (21.4); 5 years (26.0) and 7 year (39.5), while estimates from two registry-based studies were as follows 1 year (7.0, 10.4); 3 year (10.0, 15.0); 5 years (19.5, 15.0) and 7 year (22.9) [Figures 2].

### Incidence rates of dementia after ischemic stroke (per 100 person-years)

The overall proportion of dementia after ischemic stroke was 23% (18%, 29%). Reported estimates from hospital-based studies were 10% higher than those reported from registries (25% and 15%) [Figures 3-5].

**Figure 3.**
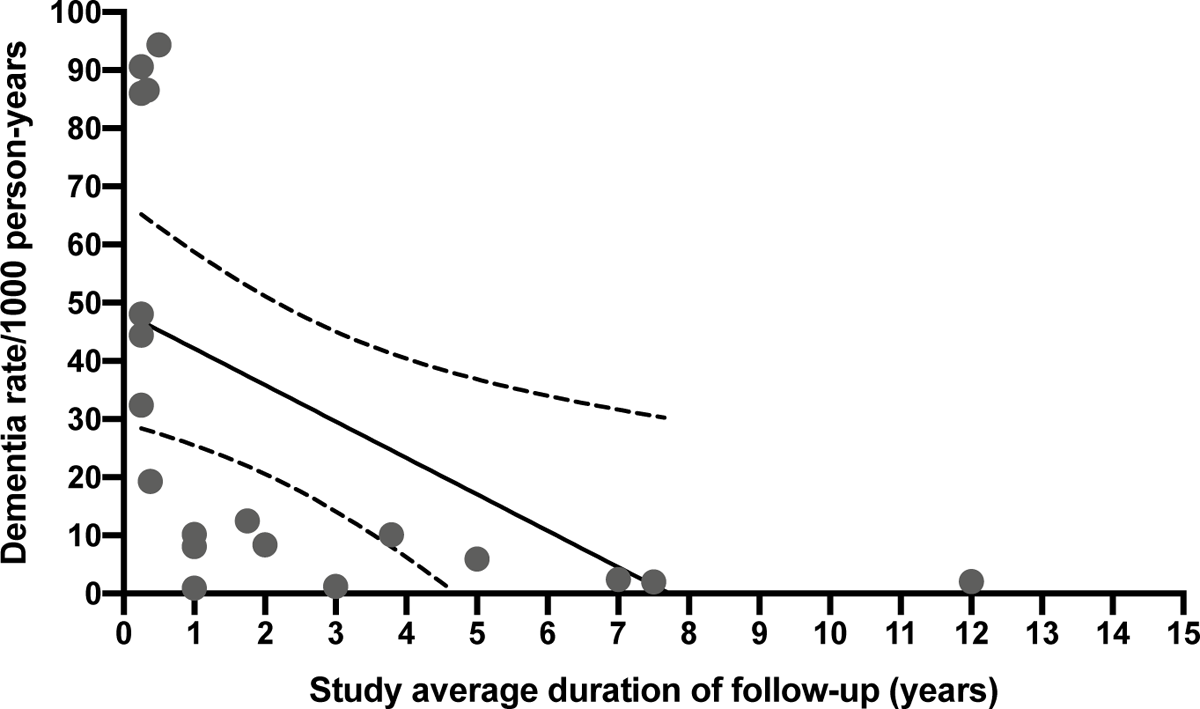
Incidence rates of dementia after ischemic stroke

**Figure 4a.**
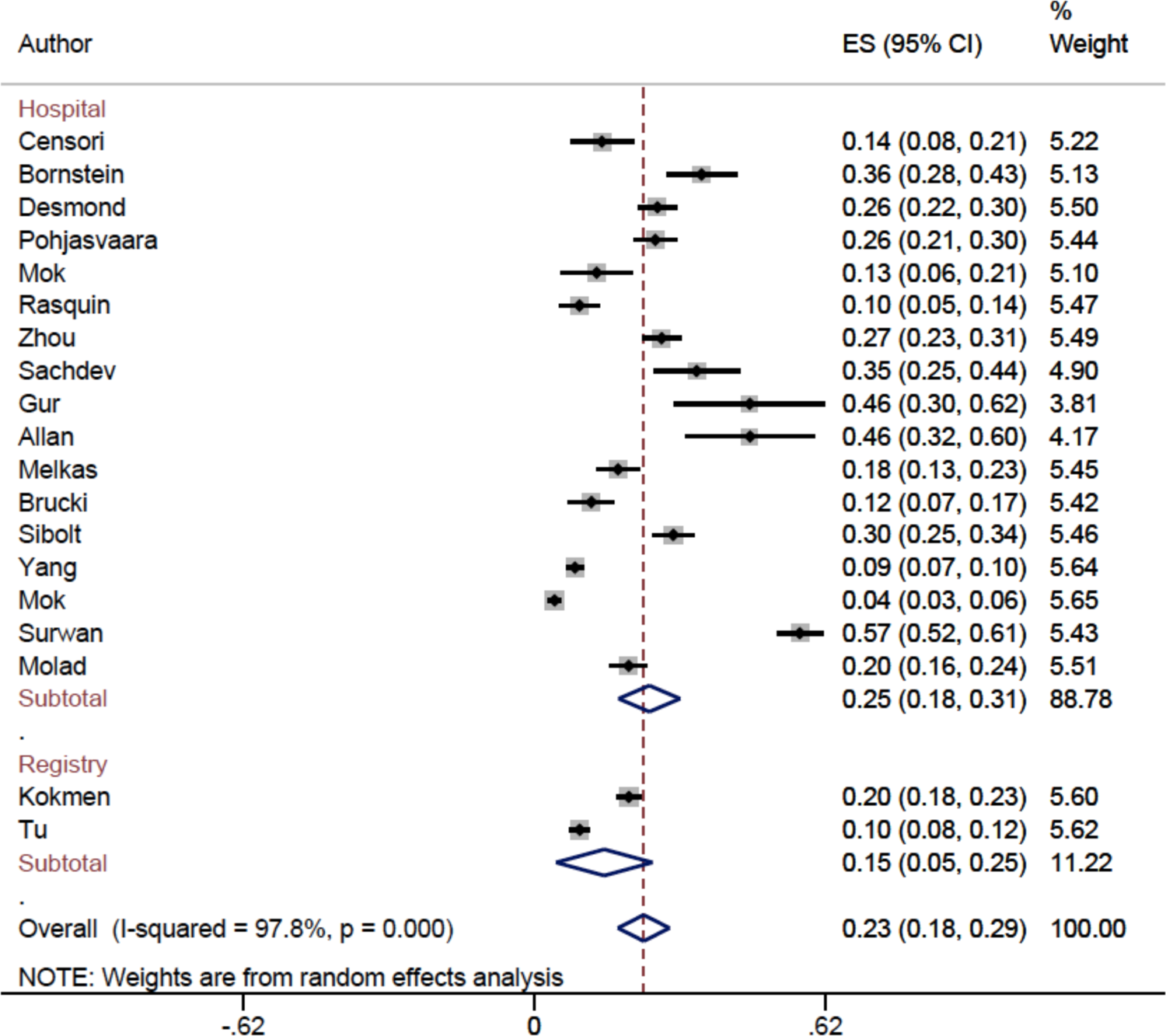
Overall proportion of dementia after ischemic stroke

**Figure 4b.**
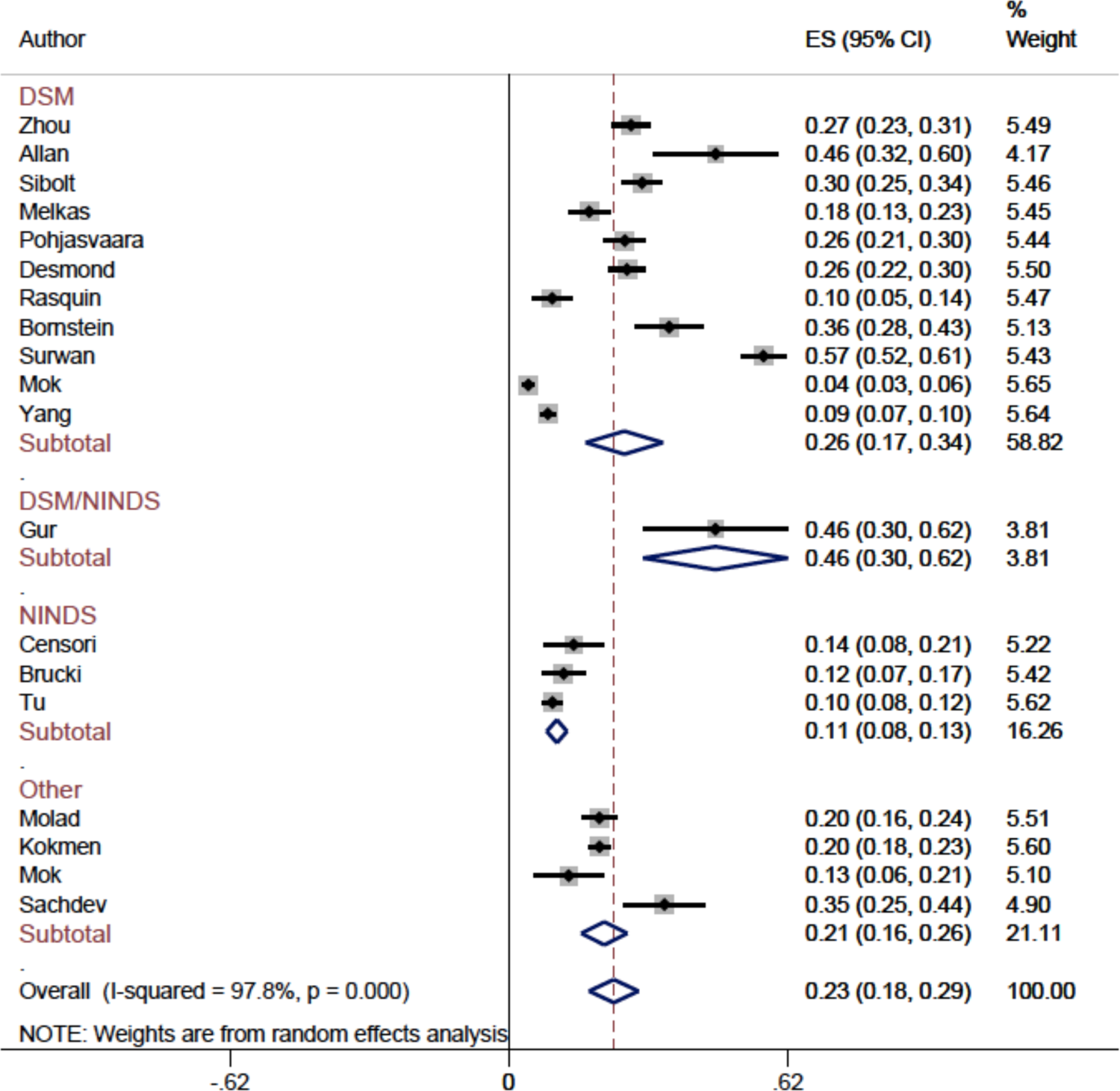
Proportion of dementia after ischemic stroke by method of diagnosis

**Figure 5a.**
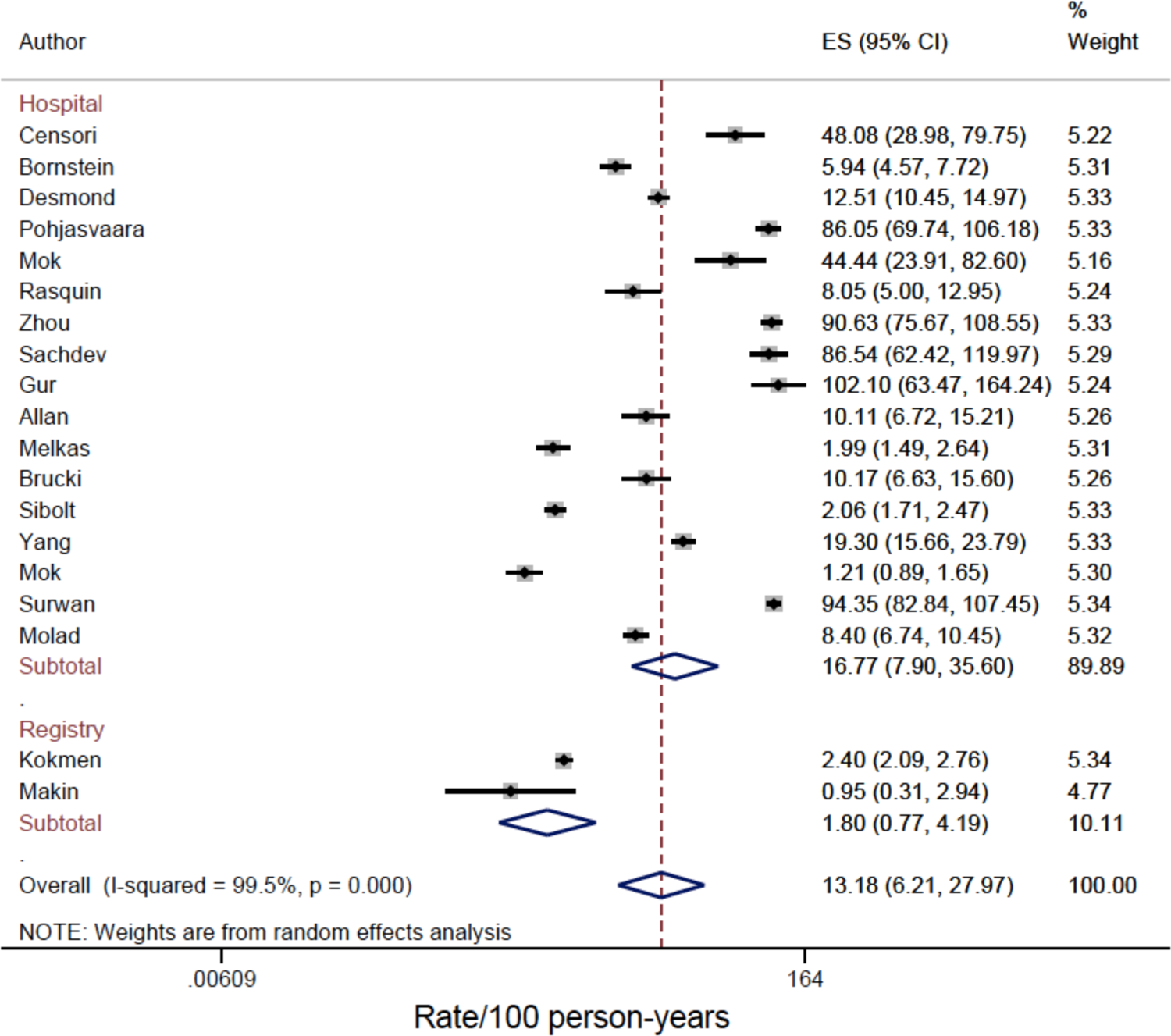
Incidence rates per 100 person-years of dementia after ischemic stroke by setting

**Figure 5b.**
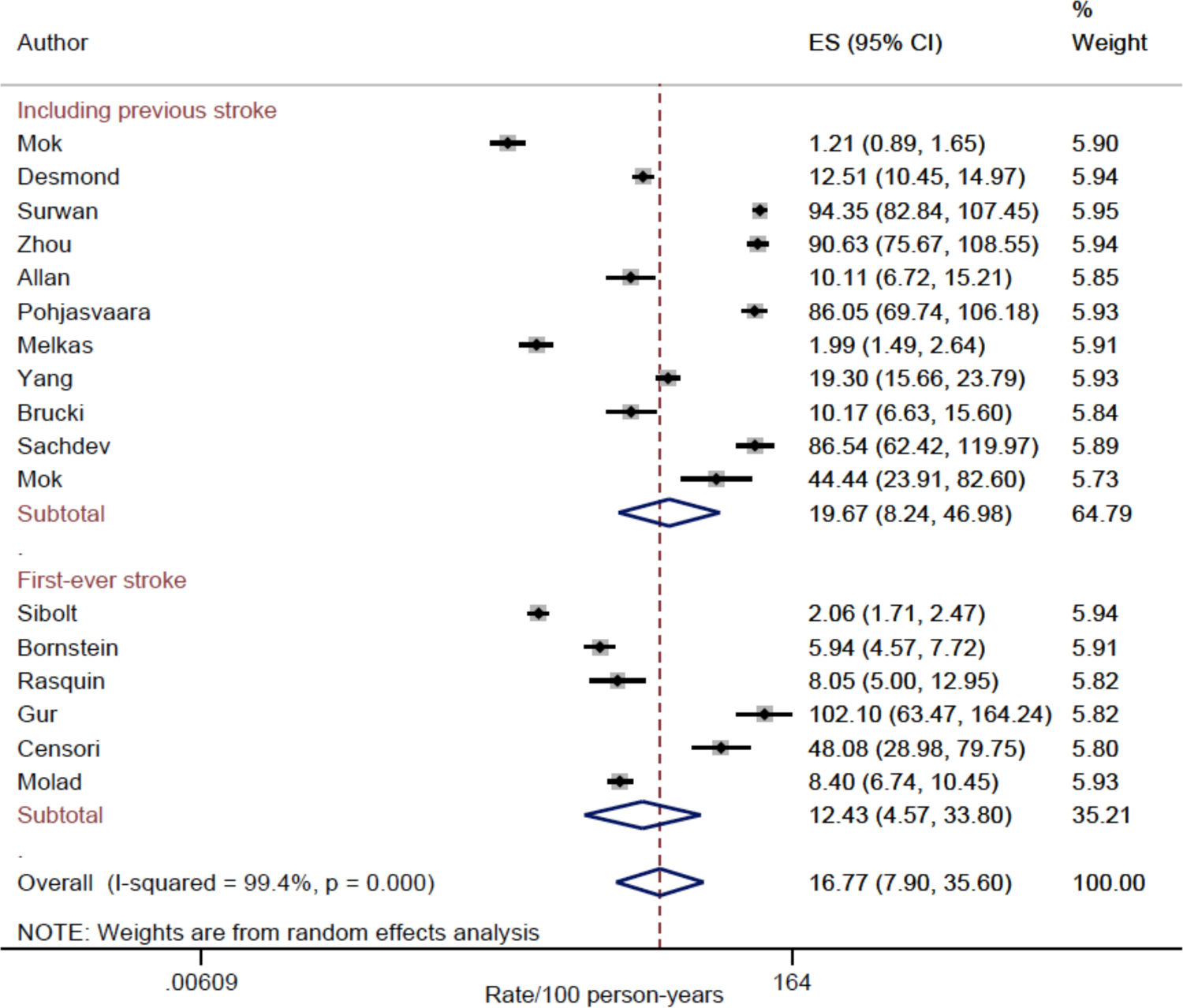
Incidence rates of dementia after ischemic stroke by presence of prior stroke

**Figure 5c.**
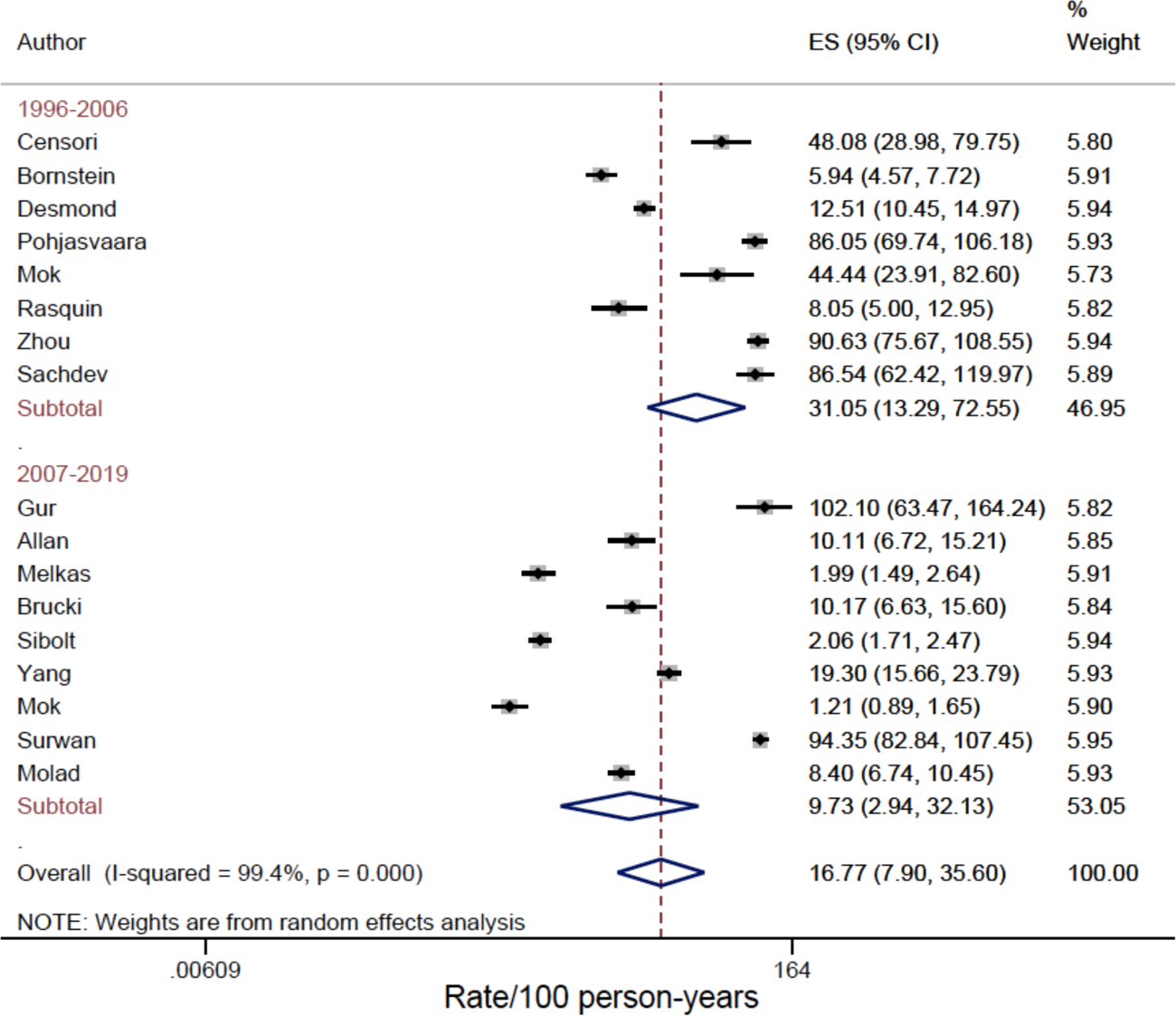
Incidence rates of dementia after ischemic stroke by study period

The overall rate of ischemic stroke after dementia was 13.2 (6.2, 28.0) per 100 person-years. In stratified analyses the incidence rates of dementia post-ischemic stroke from hospital-based studies were 16.7 per 100 person-years compared to 1.8 per 100 person-years from registry-based studies. The overall rate among those with previous strokes was 19.7 (8.2, 46.9) compared to 12.4 (4.6, 33.8) among those with first-ever stroke. The incidence rates in the first decade between 1996-2006 was 31.1 (13.3, 72.6), thus almost two-folds higher than those reported in the second decade between 2007-2019, 9.73 (2.9, 32.1) [Figure 5].

### Predictors of post-ischemic stroke dementia Demographics and other general factors

#### Gender

The overall estimated proportion of female cases was 50% and the estimated range of averages of dementia cases among females per study was between 35% and 77% [table 2]. The pooled odds ratio was 1.2 (95% CI 1.0, 1.4) [Table 3, Supplementary figure 1].

**Table 2.** Summary characteristics of individuals with and without dementia after ischemic stroke

| Characteristic | Dementia |  |  |  | No dementia |  |  |
| --- | --- | --- | --- | --- | --- | --- | --- |
|  | Study, n | mean | min | max | mean | min | max |
| Demographics |  |  |  |  |  |  |  |
| Female | 14 | 50 | 35 | 77 | 42 | 31 | 55 |
| Education, years | 5 | 7 | 3 | 11 | 6 | 4 | 11 |
| Handiness, right | 2 | 94 | 92 | 97 | 92 | 89 | 95 |
| APOE4 | 2 | 13 | 7 | 19 | 55 | 17 | 93 |
| Clinical History |  |  |  |  |  |  |  |
| HTN | 9 | 69 | 47 | 88 | 57 | 24 | 74 |
| DM | 10 | 43 | 17 | 76 | 33 | 12 | 88 |
| Smoking | 11 | 35 | 16 | 55 | 40 | 16 | 60 |
| Ischemic heart disease | 3 | 24 | 12 | 33 | 13 | 8 | 18 |
| Atrial Fibrillation | 7 | 25 | 10 | 40 | 14 | 8 | 17 |
| MI | 3 | 16 | 13 | 22 | 18 | 17 | 19 |
| Angina | 2 | 21 | 19 | 23 | 19 | 17 | 21 |
| Heart failure | 5 | 21 | 3 | 59 | 24 | 3 | 86 |
| Hypercholesterolemia | 7 | 41 | 20 | 68 | 47 | 24 | 73 |
| Alcohol | 4 | 18 | 0 | 44 | 20 | 6 | 50 |
| Previous stroke | 5 | 28 | 17 | 38 | 17 | 13 | 21 |
| Previous TIA | 5 | 12 | 2 | 25 | 10 | 3 | 17 |
| Location |  |  |  |  |  |  |  |
| Dominant hemisphere | 2 | 46 | 12 | 80 | 28 | 6 | 51 |
| Hemisphere lesion | 2 | 87 | 81 | 93 | 68 | 67 | 70 |
| MCA | 2 | 73 | 53 | 93 | 54 | 48 | 60 |
| Veretebrobasilar | 2 | 13 | 8 | 18 | 23 | 13 | 32 |
| Chronic brain changes |  |  |  |  |  |  |  |
| No. of old infarcts, mean | 2 | 5 | 3 | 8 | 3 | 2 | 6 |
| Presence of MTLA | 2 | 60 | 50 | 69 | 28.5 | 28 | 29 |
| Presence of global atrophy | 2 | 42 | 28 | 56 | 22 | 22 | 22 |
| Stroke mechanism |  |  |  |  |  |  |  |
| Atherosclerosis | 5 | 46 | 23 | 81 | 62 | 18 | 90 |
| Embolism | 5 | 20 | 15 | 24 | 13 | 6 | 20 |
| Lacunar | 3 | 33 | 22 | 41 | 36 | 32 | 38 |
| TIA | 2 | 7.4 | 7 | 8 | 15 | 15 | 15 |
| ICH | 2 | 10 | 5 | 16 | 6 | 6 | 6 |
| Unknown/other | 3 | 13 | 5 | 24 | 12 | 3 | 24 |

**Table 3.**
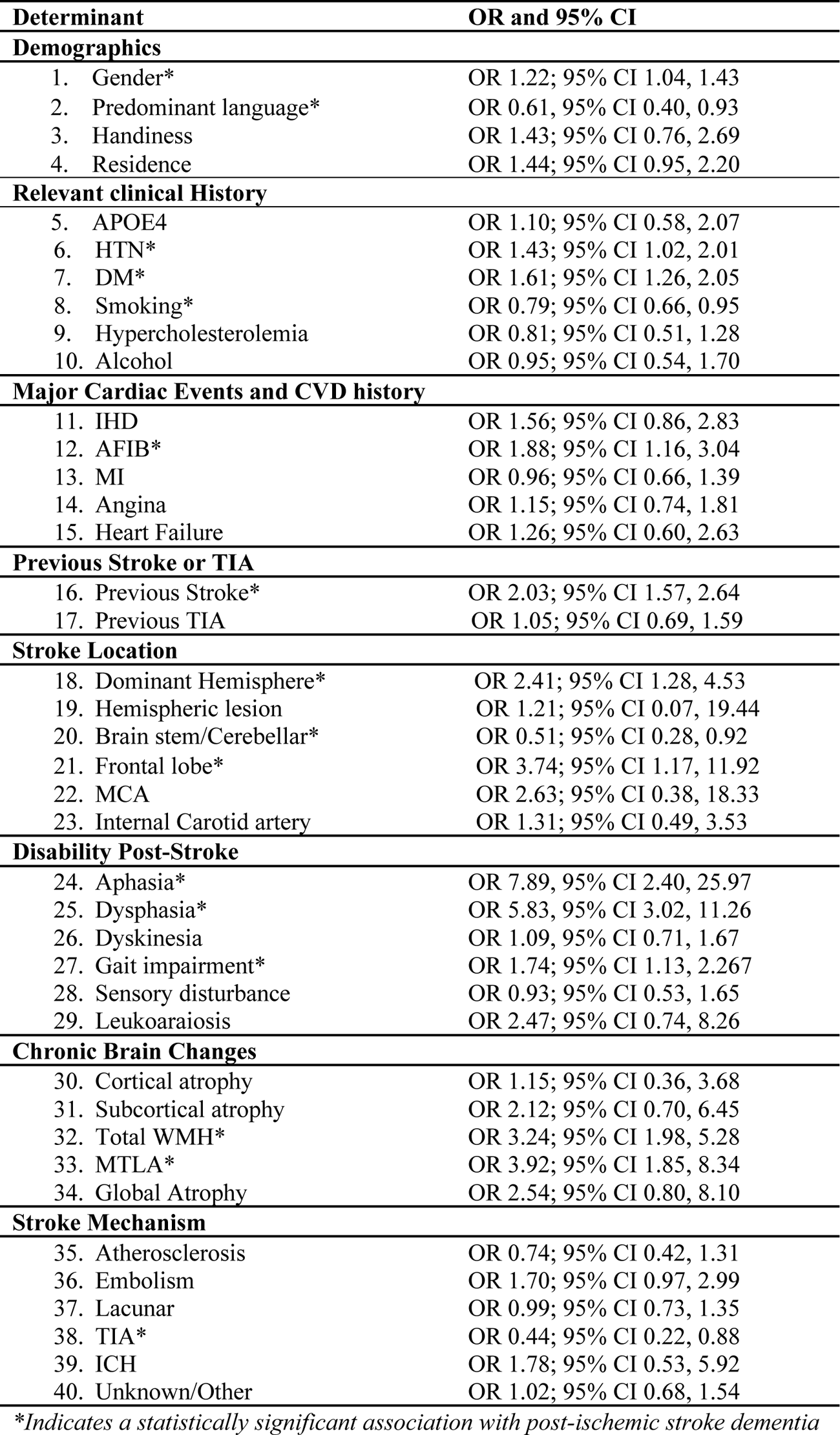
Factors associated with post-ischemic stroke dementia risk (OR**, 95% CI**

#### Right handiness

The overall estimated proportion of cases was 94% and range between 92% and 97% [table 2]. The pooled odds ratio was 1.4 (95% CI 0.8, 2.7) [Table 3, Supplementary figure 1].

### Relevant clinical history

#### APOE4

Molad et al. showed a non-significant odds ratio for post-ischemic stroke dementia by APOE4 versus no APOE4 (OR 1.1; 95% CI 0.6, 2.1) [Table 3, Supplementary figure 2].

#### Hypertension

The overall estimated proportion of dementia cases was 69% and range was between 47% and 88% [table 2]. The pooled odds ratio was (OR 1.4; 95% CI 11.0, 2.0) [Table 3, Supplementary figure 2].

#### Diabetes mellitus

The overall estimated proportion of cases was 43% and range was between 17% and 76% [table 2]. The pooled odds ratio was (OR 1.6; 95% CI 1.3, 2.1) [Table 3, Supplementary figure 2].

#### Smoking

The overall estimated proportion of cases was 35% and range was between 16% and 55% [table 2]. The pooled odds ratio was (OR 0.8; 95% CI 0.7, 0.9) [Table 3, Supplementary figure 2].

#### Hypercholesterolemia

The overall estimated proportion of cases was 41% and range was between 20% and 68% [table 2]. The pooled odds ratio was (OR 0.8; 95% CI 0.5, 1.3) [Table 3, Supplementary figure 2].

#### Alcohol

The overall estimated proportion of cases was 18% and range was between 0% and 44% [table 2]. The pooled odds ratio was (OR 0.9; 95% CI 0.5, 1.7) [Table 3, Supplementary figure 2].

### Major cardiac events

#### Ischemic heart disease

The overall estimated proportion of cases was 24% and range was between 12% and 33% [table 2]. The pooled odds ratio was (OR 1.6; 95% CI 0.8, 2.8) [Table 3, Supplementary figure 3].

#### Atrial fibrillation

The overall estimated proportion of cases was 25% and range was between 10% and 40% [table 2]. The pooled odds ratio was (OR 1.8; 95% CI 1.2, 3.0) [Table 3, Supplementary figure 3].

#### Myocardial infarction

The overall estimated proportion of cases was 16% and range was between 13% and 22% [table 2]. The pooled odds ratio was (OR 0.8; 95% CI 0.5, 1.3) [Table 3, Supplementary figure 3].

#### Angina pectoris

The overall estimated proportion of cases was 21% and range was between 19% and 23% [table 2]. The pooled odds ratio was (OR 1.2; 95% CI 0.7, 1.8) [Table 3, Supplementary figure 3].

#### Heart failure

The overall estimated proportion of cases was 21% and range was between 3% and 59% [table 2]. The pooled odds ratio was (OR 1.3; 95% CI 0.6, 2.6) [Table 3, Supplementary figure 3].

### Previous stroke or history transient ischemic attack (TIA)

Previous stroke was associated with increased odds of dementia after ischemic stroke (OR 2.0; 95% CI 1.6, 2.6, I-squared 5.1%, p=0.4), history of TIA was not associated with higher odds of post-stroke dementia (OR 1.0; 95% CI 0.7, 1.6, I-squared 13%, p=0.3) [Table 3, Supplementary figure 4].

### Stroke location

#### Dominant hemisphere

The estimated proportion of cases was 46% and range between 12% and 80% [table 2]. The pooled odds ratio was 2.4 (95% CI 1.3, 4.5) [Table 3, Supplementary figure 5].

#### Hemispheric lesion

The estimated proportion of cases with hemispheric lesion was 87% and range between 81% and 93% [Table 2]. The pooled odds ratio was 1.2 (95% CI 0.1, 19.4) [Table 3, Supplementary figure 5].

#### Middle cerebral artery (MCA)

The estimated proportion of cases was 73% and range between 53% and 93% [Table 2]. The pooled odds ratio was 2.6 (95% CI 0.4, 18.3) [Table 3, Supplementary figure 5].

#### Internal carotid artery

The estimated proportion of cases was 5.0 % compared to 3.8% for non-cases [table 2]. The pooled odds ratio was 1.3 (95% CI 0.5, 3.5) [Table 3, Supplementary figure 5].

#### Brain stem/Cerebellar

The estimated proportion of cases was 18.5% compared to 32.9% [table 2]. The pooled odds ratio was 0.5 (95% CI 0.3, 0.9) [Table 3, Supplementary figure 5].

#### Frontal lobe

The estimated proportion of cases was 66.7% for cases and 35% for non-cases [table 2]. The pooled odds ratio was 3.7 (95% CI 1.2, 11.9) [Table 3, Supplementary figure 5].

### Disability post-stroke

#### Aphasia

One study by Censori et al. showed higher odds of post stroke dementia for aphasia (OR 7.8, 95% CI 2.4, 25.9) (Table 3, Supplementary figure 6).

#### Dysphasia, gait impairment

Zhou et al. demonstrated a higher risk for post-ischemic stroke dementia according to presence of dysphasia (OR 5.8, 95% CI 3.0, 11.3) and among those who developed gait impairment due to stroke 1.7 (95% CI 1.1, 2.3) [Table 3, Supplementary figure 6].

#### Dyskinesia and sensory disturbance

The study by Zhou et al showed a non-significant association with post-ischemic stroke dementia according to presence of dyskinesia (OR 1.1, 95% CI 0.7, 1.7) or sensory disturbance 0.9 (95% CI 0.5, 1.6) (Table 3, Supplementary figure 6).

#### Leukoaraiosis

Censori et al showed a non-significant association between leukoaraiosis and post-ischemic stroke dementia risk 2.5 (95% CI 0.7, 8.3) [Table 3, Supplementary figure 6].

### Chronic brain changes

#### Cortical atrophy

One study by Censori et al showed non-significant associations between post-ischemic stroke dementia risk and presence of cortical atrophy 1.2 (95% CI 0.4, 3.7) or subcortical atrophy 2.1 (95% CI 0.7, 6.5) [Table 3, Supplementary figure 6].

#### Total white matter hyperintensities (WMH)

The estimated proportion of cases was 9.5% for cases and 6.4% for non-cases [table 2]. The pooled odds ratio was 3.2 (95% CI 1.9, 5.3) [Table 3, Supplementary figure 6].

#### Medial temporal lobe atrophy (MTLA)

The estimated proportion of cases was 60% and range between 50% and 69% [table 2]. The pooled odds ratio was 3.9 (95% CI 1.8, 8.3) [Table 3, Supplementary figure 6].

#### Global atrophy

The estimated proportion of cases was 42% and range between 28% and 56% [table 2]. The pooled odds ratio was 2.5 (95% CI 0.8, 8.1) [Table 3, Supplementary figure 6].

### Stroke mechanism

#### Atherosclerosis

The estimated proportion of cases was 46% and range between 23% and 81% [table 2]. The pooled odds ratio was 0.7 (95% CI 0.4, 1.3) [Table 3, Supplementary figure 7].

#### Embolism

The estimated proportion of cases was 20% and range between 15% and 24% [table 2]. The pooled odds ratio was 1.7 (95% CI 0.9, 2.9) [Table 3, Supplementary figure 7].

#### Lacunar stroke

The estimated proportion of cases was 33% and range between 22% and 41% [table 2]. The pooled odds ratio was 0.9 (95% CI 0.7, 1.4) [Table 3, Supplementary figure 7].

#### Transient ischemic attacks

The estimated proportion of cases was 7.4% and range between 7 % and 8% [table 2]. The pooled odds ratio was 0.4 (95% CI 0.2, 0.8) [Table 3, Supplementary figure 7].

#### Intracerebral hemorrhage

The estimated proportion of cases was 10% and range between 5% and 16% [table 2]. The pooled odds ratio was 1.8 (95% CI 0.5, 5.9) [Table 3, Supplementary figure 7].

## DISCUSSION

In the present report, we synthesized the literature to summarize incidence rates and predictors of post-ischemic stroke dementia. Our analysis included 21 studies representing 55,183 patients from studies published between 1996 up to 2019 in the Americas, Europe, Asia and Australia.

We analyzed 40 predictors of dementia after ischemic stroke including demographics, relevant clinical history, major cardiac events, previous TIA or stroke, stroke location, disability after stroke, chronic brain changes and stroke mechanism. We have also assessed study related factors including study setting, period and method of dementia diagnosis.

Incidence rates of dementia were much higher in studies with very short follow-up, those conducted in a clinic or hospital setting compared to population-based studies and among those where DSM alone was the main method of diagnosis compared to NINDS alone or combined with DSM. There was almost 33% difference in incidence rates of dementia in the later study period (2007 – 2019) compared to (1996 – 2006). Incidence rates of dementia after ischemic stroke were 1.5 times higher in the presence of stroke history as opposed to first-time occurrence. Predictors that were strongly associated with dementia occurrence after ischemic stroke were female gender, hypertension, diabetes mellitus, atrial fibrillation, previous stroke, presence of stroke lesion in (dominant hemisphere, brain stem/cerebellum or frontal lobe), presence of aphasia, dysphasia, gait impairment, presence of white matter hyperintensities, medial temporal lobe atrophy and TIA as the leading mechanism for ischemic stroke.

Progression to dementia among stroke survivors is multifactorial and could be explained within three main categories, 1) patient-related factors including demographics, 3) severity and location of the presenting ischemic stroke and 3) presence of relevant clinical comorbidities, vascular risk factors, and diffuse cerebrovascular disease. Ischemic infarcts frequently occur alongside arteriosclerotic small vessels disease, which may further contribute to cerebral atrophy and cognitive decline^27–29^. Ischemic lesions of the left hemisphere correlate with intellectual deterioration. It has been suggested that the left hemisphere is responsible for both language and generalized cognitive function^17, 30, 31^. Further, large lesions in the dominant hemisphere in the middle cerebral artery and left carotid artery territories are strongly associated with dementia^17, 30–32^. Evidence also suggests the involvement of limbic structures among stroke patients and this points to the importance of the medial frontal and medial temporal lobes in memory^17^. Other important factors include presence of a previous stroke that has been independently associated with risk of dementia^18^. In our review, studies that included patients with recurrent stroke reported generally higher incidence rates of dementia.

Individual specific characteristics are key in understanding progression to dementia after ischemic stroke. Older age, years of education and non-white race have been associated with occurrence of dementia among stroke patients in general. In our study, age ranged between 65 and 80, thus likely representing a population of patients who are at higher risk compared to the general population. Previous studies were inconsistent regarding the role of sex^18, 33, 34^. Our results suggest women to have higher odds of dementia after ischemic stroke.

Chronic hypoperfusion is an important underlying mechanism through which cardiovascular risk factors could contribute to cognitive decline and dementia in the setting of ischemia. These changes result in leukoaraiosis with subsequent functional consequences and impairment of emboli clearance from the brain^35–37^. Our results confirm that patients with atrial fibrillation represent a very high-risk group for dementia after ischemic stroke. This is consistent with the literature on increased risk of dementia after any stroke^18, 38^. Similarly, the presence of diabetes mellitus has been associated with high risk of dementia through indirect effects on cerebral blood flow that are possibly linked to disruption in autoregulation or indirectly as a risk factor for silent brain infarction^39^.

Our study has several limitations. First, it was not possible to conduct a pooled analysis for all factors of interest due to limited data by stroke-subtype in individual studies. However, to account for this limitation we leveraged data from studies that reported case counts to calculate the odds ratios for predictors of interest. Further, the included studies were heterogeneous in terms of follow-up duration after stroke onset. Therefore, we restricted the combined analysis to studies with more than 6 months of follow-up to minimize reverse causality. Lastly, our results are mainly among patients between 65 and 80 years old and thus cannot be extrapolated to younger or older ischemic stroke patients.

Through the present systematic review and meta-analysis, we assessed frequency, patient-level predictors (demographics, CVD history and major cardiac events, previous stroke and TIA, stroke location, disability post-stroke, chronic brain change and stroke mechanism) and study-level predictors of dementia after ischemic stroke (setting; hospital or registry), (methods of dementia diagnosis; DSM, NINDS or both), (first-ever stroke or recurrent stroke).

Given the variations in estimates on post-stroke dementia in the ischemic stroke population according to study-level predictors, comparing evidence from various data sources and study settings is essential. Post-ischemic stroke dementia is a distinct subtype of vascular dementia with clear differences in patient-level predictors from those of general dementia. Information collected at time of ischemic stroke onset can be collectively used to monitor dementia risk post-ischemic stroke.

## Data Availability

All data produced are available within the article with accurate and complete citations.

---

**Supplementary figure 1.**
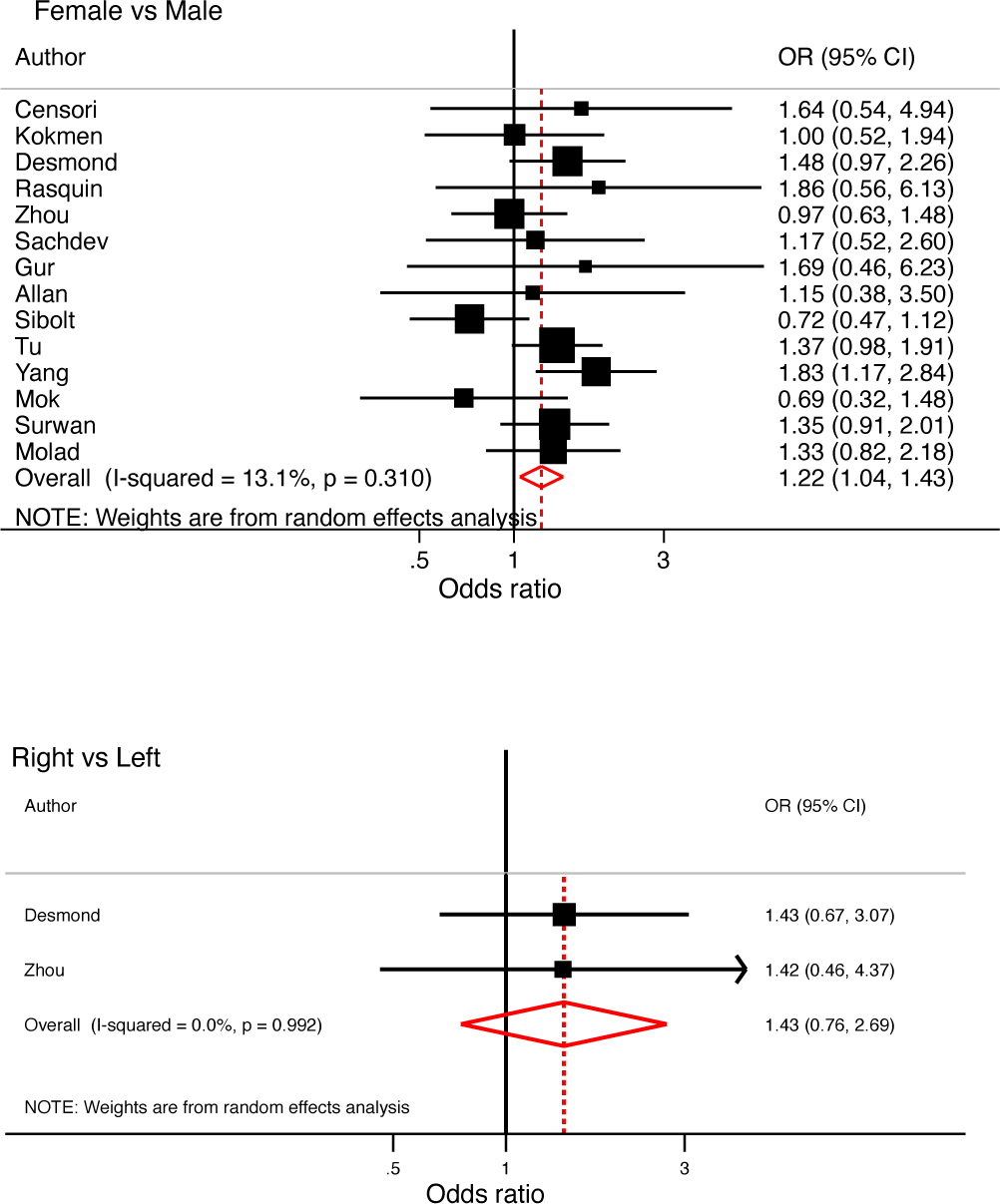
Forest plots of gender and handiness in relation to post-ischemic stroke dementia

**Supplementary figure 2.**
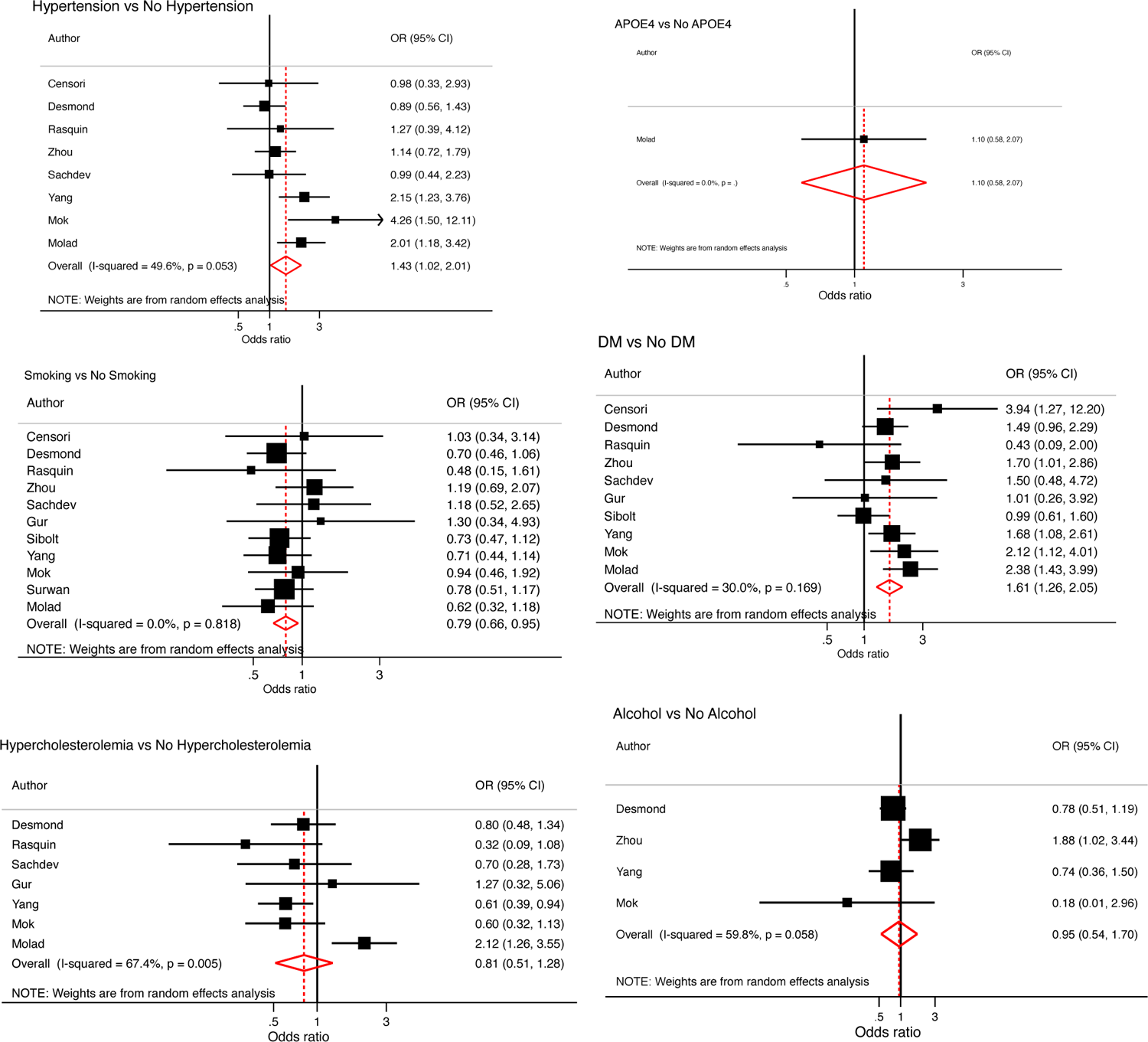
Forest plots of clinical history in relation to post-ischemic stroke dementia, a) Hypertension; b) APOE4; c) DM; d) Smoking; e) hypercholesterolemia and f) alcohol

**Supplementary figure 3.**
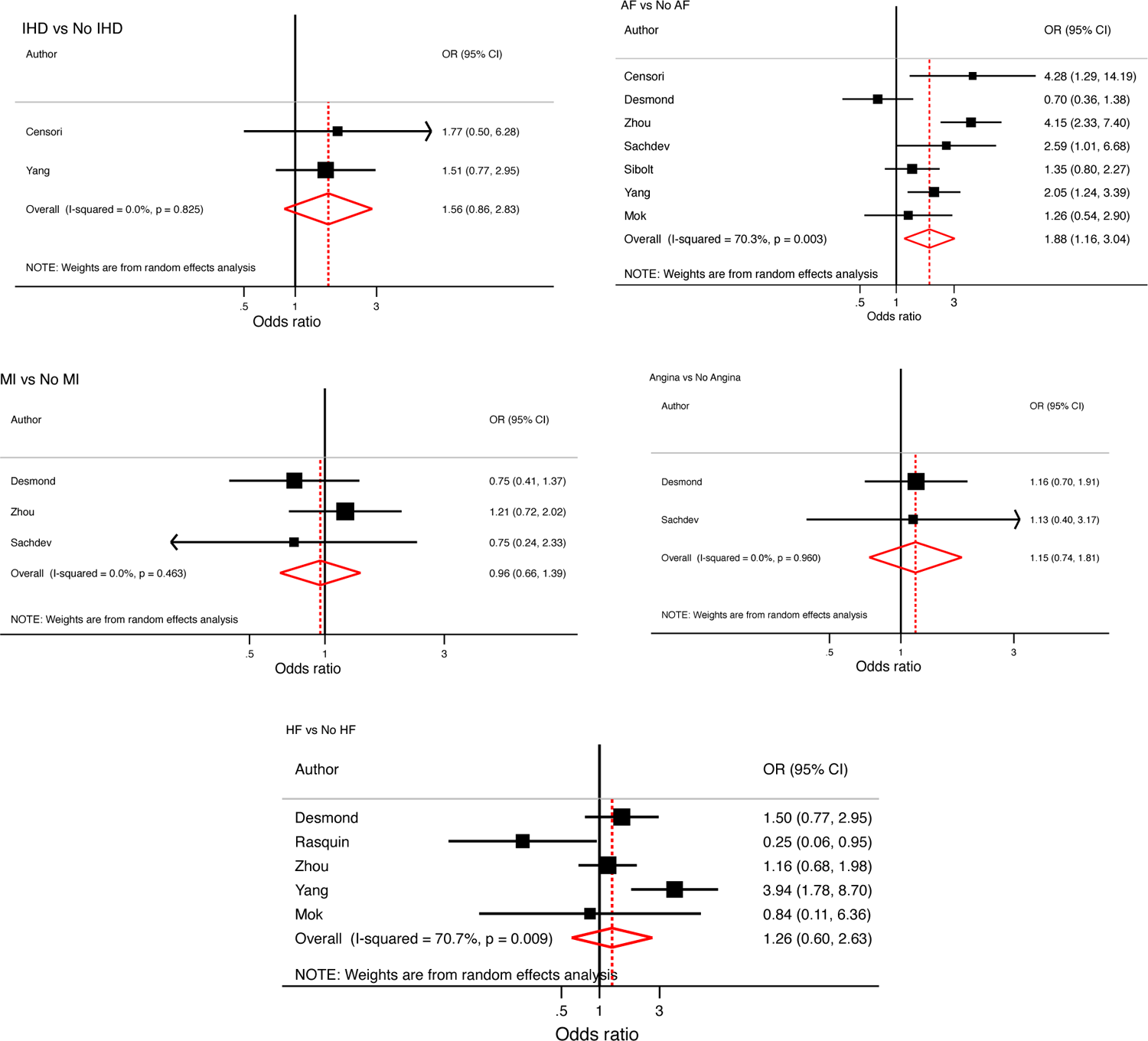
Forest plots of major cardiac events in relation to post-ischemic stroke dementia risk a) ischemic heart disease; b) Atrial fibrillation; c) Myocardial infarction; d) Angina, and e) heart failure.

**Supplementary figure 4.**
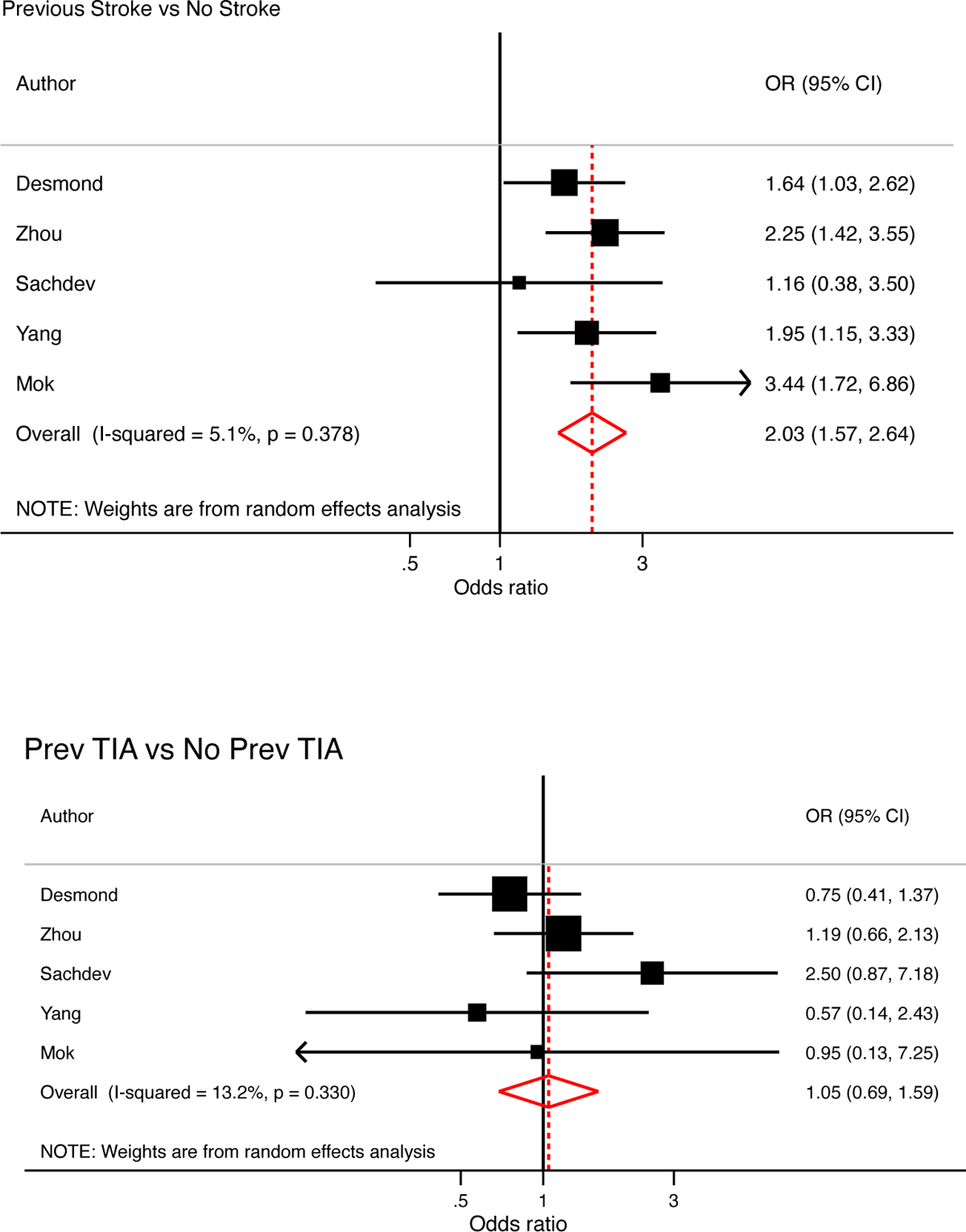
Forest plots of a) previous stroke, and b) TIA in relation to post-ischemic stroke dementia risk

**Supplementary figure 5.**
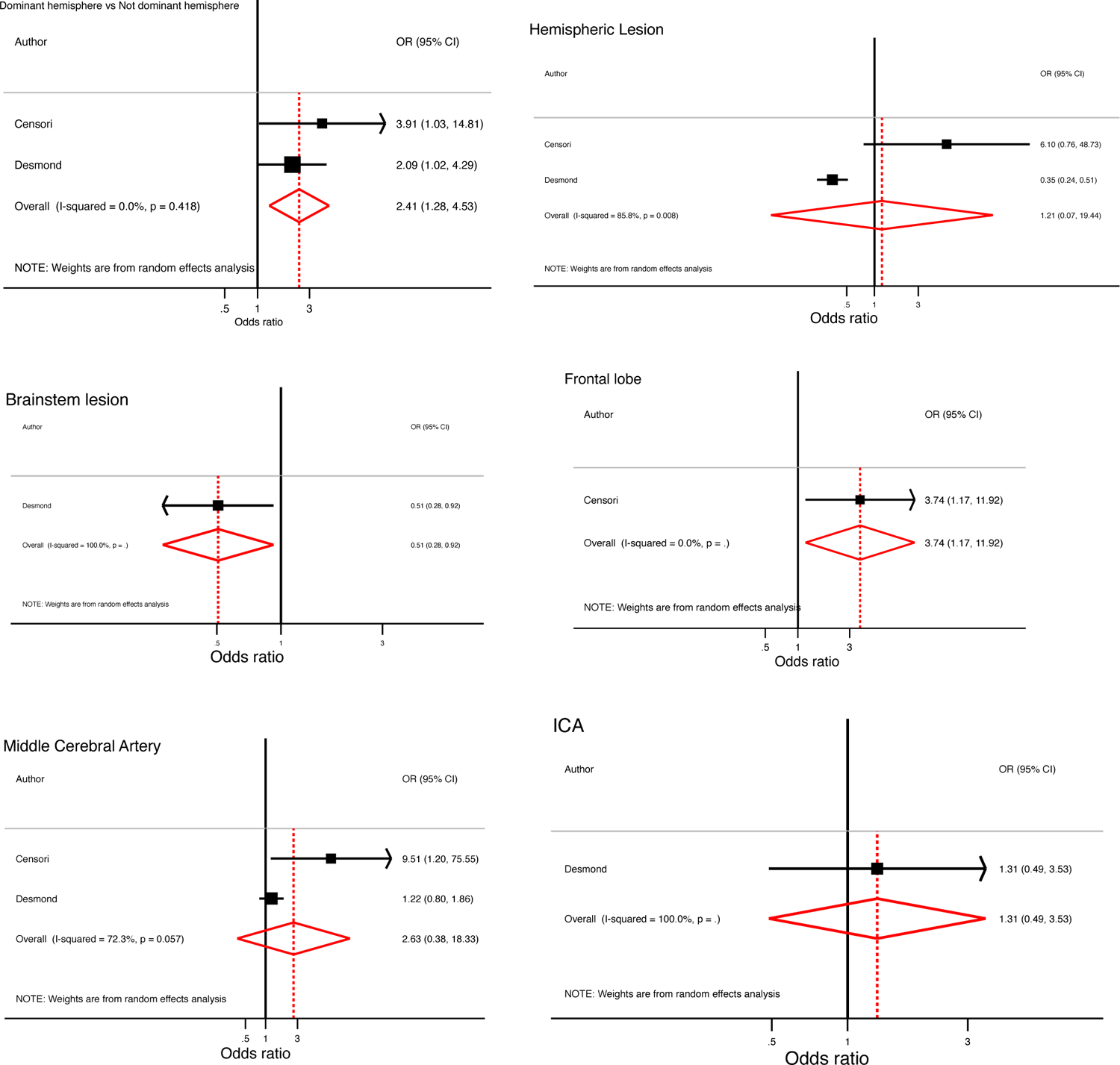
Forest plots of stroke location in relation to post-ischemic stroke dementia risk: a) dominant hemisphere; b) hemispheric lesion; c) brain stem/cerebellar; d) frontal lobe; e) MCA and f) internal carotid artery

**Supplementary figure 6.**
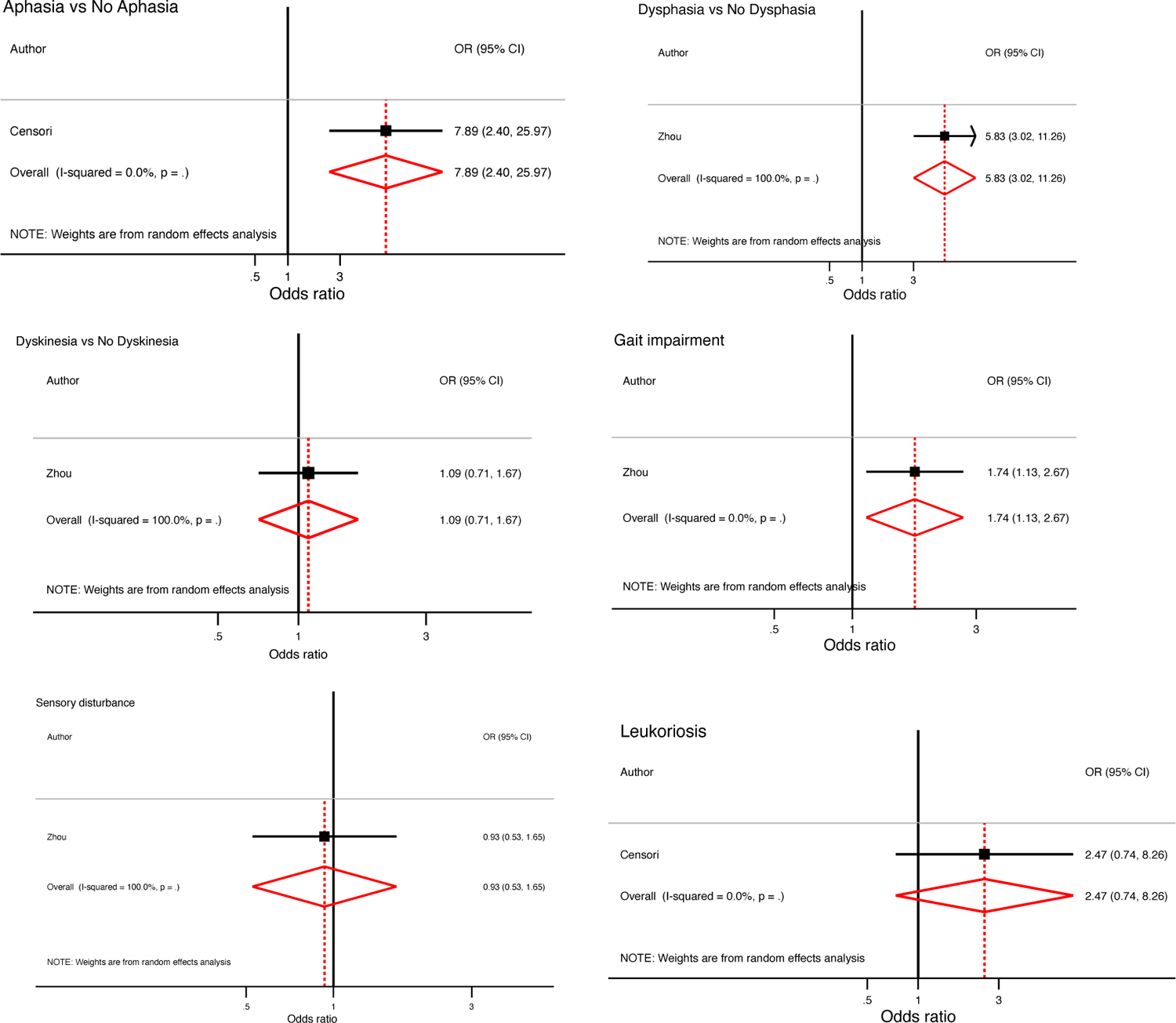
Forest plots of stroke disability-related factors in relation to post-ischemic stroke dementia risk: a) aphasia; b) dysphasia; c) dyskinesia; d) gait impairment; g) sensory disturbance and e) leukoaraiosis

**Supplementary figure 7.**
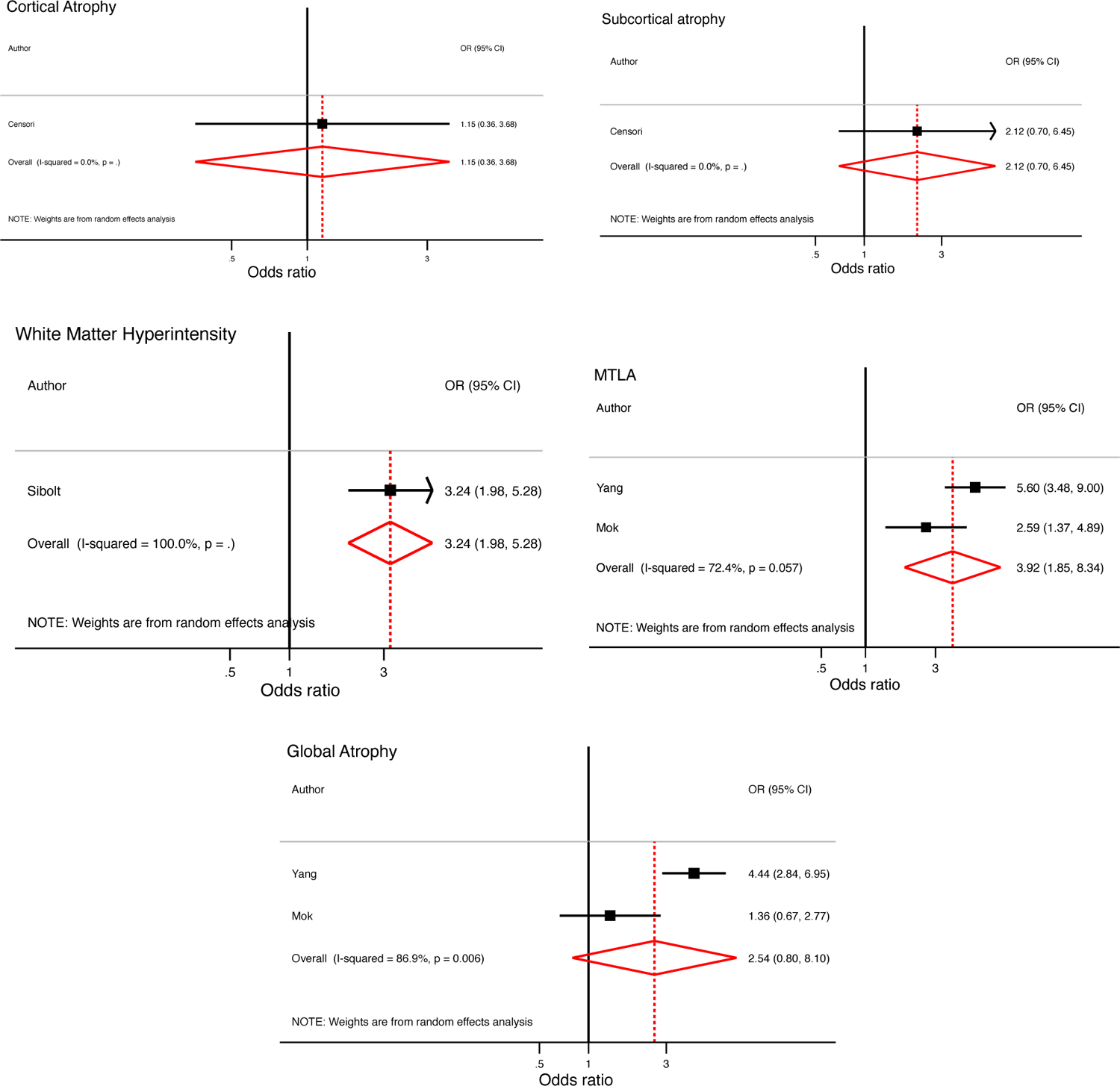
Forest plots of chronic cortical brain changes in relation to post-ischemic stroke dementia risk: a) cortical atrophy; b) subcortical atrophy; c) total WMH; d) MTLA and e) global atrophy

**Supplementary figure 8.**
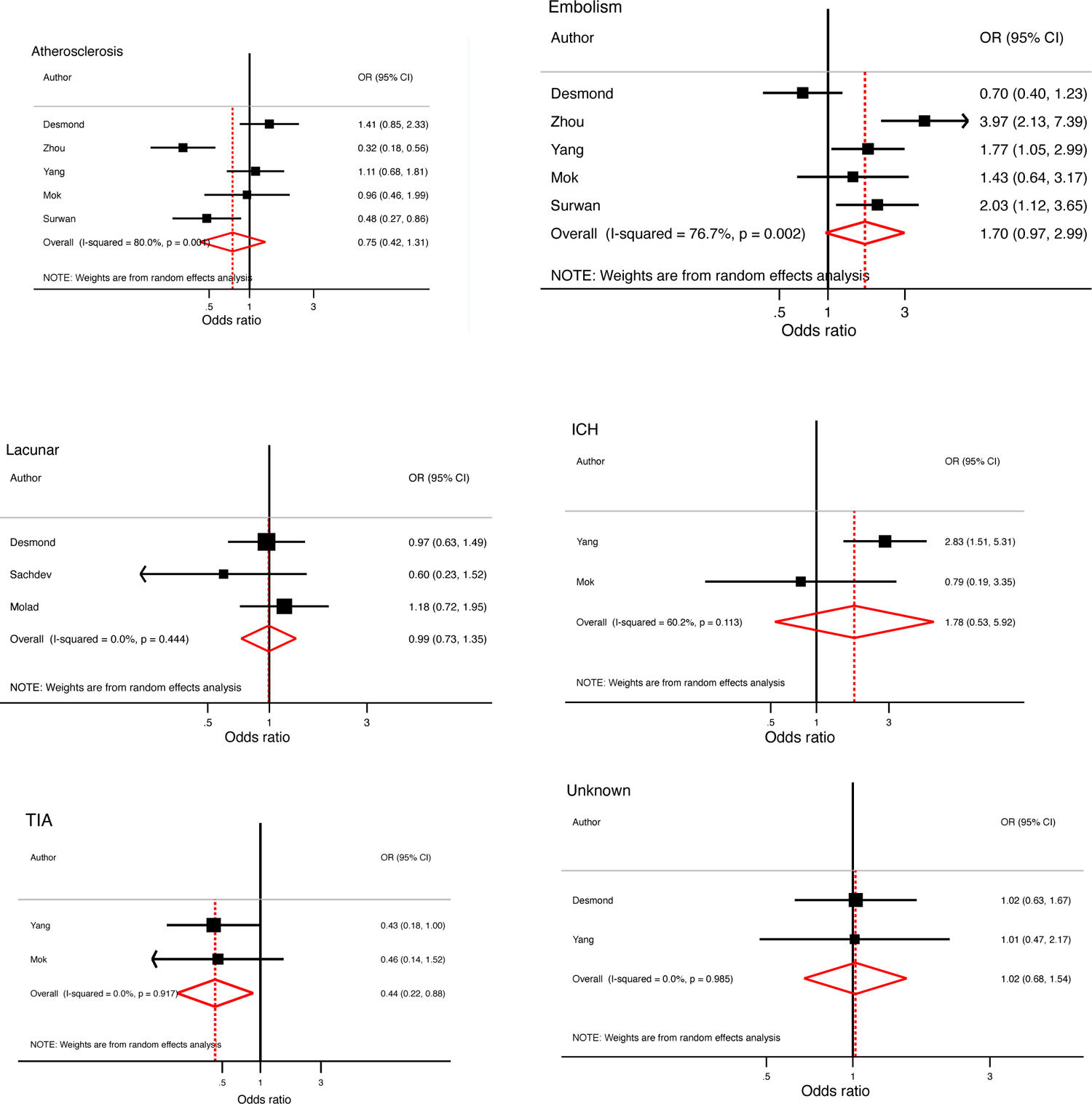
Forest plots of stroke mechanism in relation to post-ischemic stroke dementia risk: a) atherosclerosis; b) embolism; c) lacunar; d) TIA; e) ICH and f) unknown

**Supplementary table 1.**
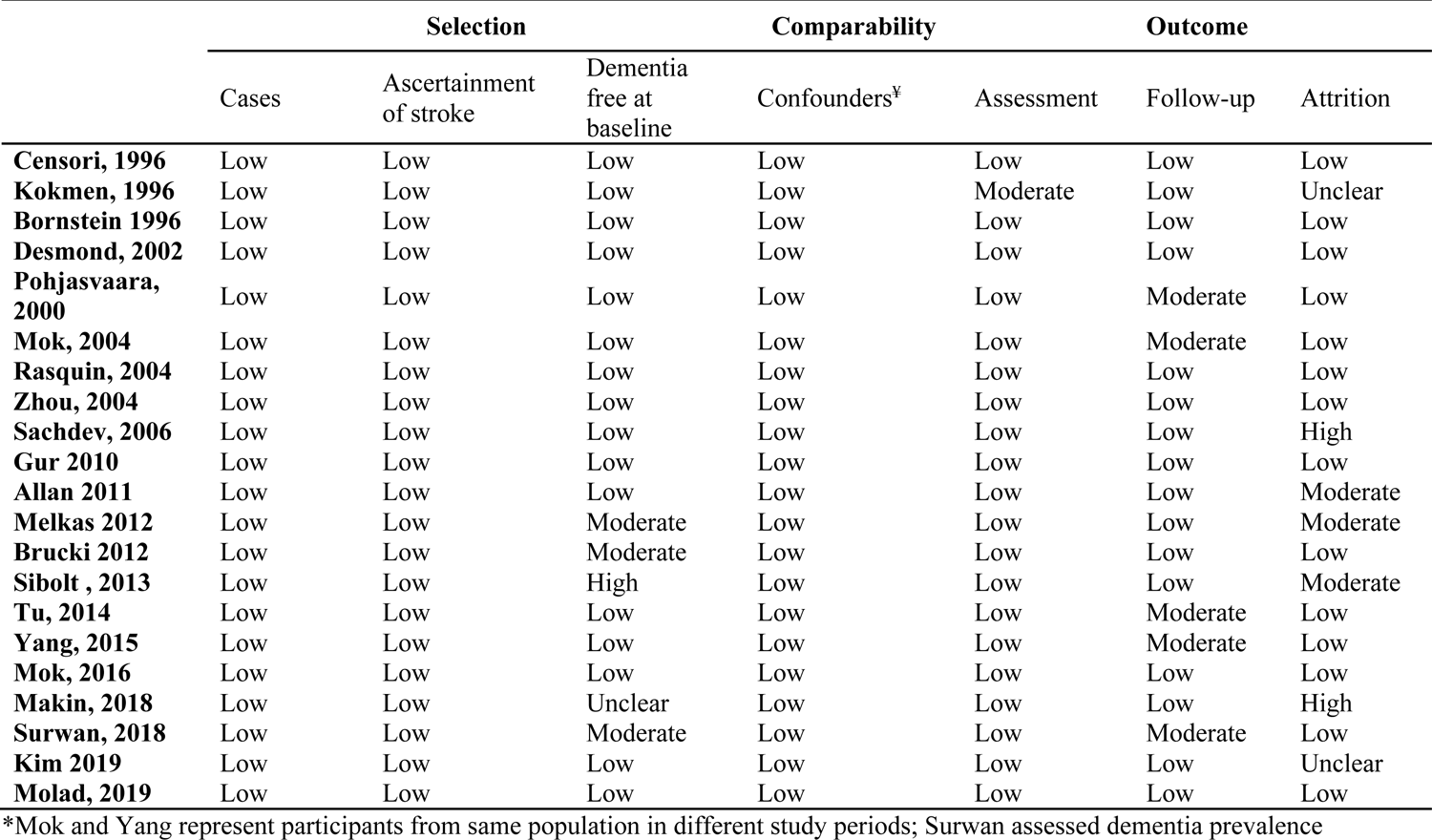
Risk of bias assessment based on Newcastle-Ottawa assessment (ranked as low, moderate or high risk)

**Supplementary table 2:** Preferred Reporting Items for Systematic reviews and Meta-Analyses (PRISMA) checklist.

| Section/topic | # | Checklist item | Reported on page # |
| --- | --- | --- | --- |
| **TITLE** |  |  |  |
| Title | 1 | Identify the report as a systematic review, meta-analysis, or both. | 1 |
| **ABSTRACT** |  |  |  |
| Structured summary | 2 | Provide a structured summary including, as applicable: background; objectives; data sources; study eligibility criteria, participants, and interventions; study appraisal and synthesis methods; results; limitations; conclusions and implications of key findings; systematic review registration number. | 2-3 |
| **INTRODUCTION** |  |  |  |
| Rationale | 3 | Describe the rationale for the review in the context of what is already known. | 4 |
| Objectives | 4 | Provide an explicit statement of questions being addressed with reference to participants, interventions, comparisons, outcomes, and study design (PICOS). | 4 |
| **METHODS** |  |  |  |
| Protocol and registration | 5 | Indicate if a review protocol exists, if and where it can be accessed (e.g., Web address), and, if available, provide registration information including registration number. | 5-6 |
| Eligibility criteria | 6 | Specify study characteristics (e.g., PICOS, length of follow-up) and report characteristics (e.g., years considered, language, publication status) used as criteria for eligibility, giving rationale. | 5-6 |
| Information sources | 7 | Describe all information sources (e.g., databases with dates of coverage, contact with study authors to identify additional studies) in the search and date last searched. | 5-6 |
| Search | 8 | Present full electronic search strategy for at least one database, including any limits used, such that it could be repeated. | 5-6 |
| Study selection | 9 | State the process for selecting studies (i.e., screening, eligibility, included in systematic review, and, if applicable, included in the meta-analysis). | 5-6 |
| Data collection process | 10 | Describe method of data extraction from reports (e.g., piloted forms, independently, in duplicate) and any processes for obtaining and confirming data from investigators. | 5-6 |
| Data items | 11 | List and define all variables for which data were sought (e.g., PICOS, funding sources) and any assumptions and simplifications made. | 5-6 |
| Risk of bias in individual studies | 12 | Describe methods used for assessing risk of bias of individual studies (including specification of whether this was done at the study or outcome level), and how this information is to be used in any data synthesis. | 5-6 |
| Summary measures | 13 | State the principal summary measures (e.g., risk ratio, difference in means). | 5-6 |
| Synthesis of results | 14 | Describe the methods of handling data and combining results of studies, if done, including measures of consistency (e.g., $I^2$) for each meta-analysis. | 5-6 |
| Risk of bias across studies | 15 | Specify any assessment of risk of bias that may affect the cumulative evidence (e.g., publication bias, selective reporting within studies). | 5-6 |
| Additional analyses | 16 | Describe methods of additional analyses (e.g., sensitivity or subgroup analyses, meta-regression), if done, indicating which were pre-specified. | 5-7 |
| **RESULTS** |  |  |  |
| Study selection | 17 | Give numbers of studies screened, assessed for eligibility, and included in the review, with reasons for exclusions at each stage, ideally with a flow diagram. | 8-15 |
| Study characteristics | 18 | For each study, present characteristics for which data were extracted (e.g., study size, PICOS, follow-up period) and provide the citations. | 8-15 |
| Risk of bias within studies | 19 | Present data on risk of bias of each study and, if available, any outcome level assessment (see item 12). | 8-15 |
| Results of individual studies | 20 | For all outcomes considered (benefits or harms), present, for each study: (a) simple summary data for each intervention group (b) effect estimates and confidence intervals, ideally with a forest plot. | 8-15 |
| Synthesis of results | 21 | Present results of each meta-analysis done, including confidence intervals and measures of consistency. | 8-15, 26-46 |
| Risk of bias across studies | 22 | Present results of any assessment of risk of bias across studies (see Item 15). | 8-15,26-46 |
| Additional analysis | 23 | Give results of additional analyses, if done (e.g., sensitivity or subgroup analyses, meta-regression [see Item 16]). | 8-15 |
| **DISCUSSION** |  |  |  |
| Summary of evidence | 24 | Summarize the main findings including the strength of evidence for each main outcome; consider their relevance to key groups (e.g., healthcare providers, users, and policy makers). | 16-19 |
| Limitations | 25 | Discuss limitations at study and outcome level (e.g., risk of bias), and at review-level (e.g., incomplete retrieval of identified research, reporting bias). | 16-19 |
| Conclusions | 26 | Provide a general interpretation of the results in the context of other evidence, and implications for future research. | 16-19 |
| **FUNDING** |  |  |  |
| Funding | 27 | Describe sources of funding for the systematic review and other support (e.g., supply of data); role of funders for the systematic review. | 2 |

From: Moher D, Liberati A, Tetzlaff J, Altman DG, The PRISMA Group (2009). Preferred Reporting Items for Systematic Reviews and Meta-Analyses: The PRISMA Statement. PLoS Med 6(7): e1000097. doi:10.1371/journal.pmed1000097

## REFERENCES

1. Benjamin EJ, Muntner P, Bittencourt MS. Heart disease and stroke statistics-2019 update: a report from the American Heart Association. Circulation. 2019;139(10):e56-e528.

2. Ivan CS, Seshadri S, Beiser A, et al. Dementia after stroke: the Framingham Study. Stroke. 2004;35(6):1264–1268.

3. Kase C, Wolf P, Kelly-Hayes M, Kannel W, Beiser A, D’Agostino R. Intellectual decline after stroke: the Framingham Study. Stroke. 1998;29(4):805–812.

4. Reitz C, Bos MJ, Hofman A, Koudstaal PJ, Breteler MM. Prestroke cognitive performance, incident stroke, and risk of dementia: the Rotterdam Study. Stroke. 2008;39(1):36–41.

5. Jin Y-P, Di Legge S, Ostbye T, Feightner JW, Hachinski V. The reciprocal risks of stroke and cognitive impairment in an elderly population. Alzheimer’s & dementia. 2006;2(3):171–178.

6. Pendlebury ST, Rothwell PM. Prevalence, incidence, and factors associated with pre-stroke and post-stroke dementia: a systematic review and meta-analysis. The Lancet Neurology. 2009;8(11):1006–1018.

7. Kuźma E, Lourida I, Moore SF, Levine DA, Ukoumunne OC, Llewellyn DJ. Stroke and dementia risk: A systematic review and meta-analysis. Alzheimer’s & Dementia. 2018;14(11):1416–1426.

8. Liberati A, Altman DG, Tetzlaff J, et al. The PRISMA statement for reporting systematic reviews and meta-analyses of studies that evaluate health care interventions: explanation and elaboration. PLoS medicine. 2009;6(7):e1000100.

9. Higgins JP, Altman DG, Gøtzsche PC, et al. The Cochrane Collaboration’s tool for assessing risk of bias in randomised trials. Bmj. 2011;343:d5928.

10. Wells G. Shea. B., O’Connell. D., Peterson J, Welch V, Losos M, et al. The Newcastle Ottawa Scale (NOS) for assessing the quality of nonrandomized studies in meta-analyses. In:2011.

11. Harris RJ, Deeks JJ, Altman DG, Bradburn MJ, Harbord RM, Sterne JA. Metan: fixed- and random-effects meta-analysis. The Stata Journal. 2008;8(1):3–28.

12. Ioannidis JP, Patsopoulos NA, Evangelou E. Uncertainty in heterogeneity estimates in meta-analyses. Bmj. 2007;335(7626):914-916.

13. Sterne JA, Egger M, Smith GD. Investigating and dealing with publication and other biases in meta-analysis. Bmj. 2001;323(7304):101-105.

14. Censori B, Manara O, Agostinis C, et al. Dementia after first stroke. Stroke. 1996;27(7):1205–1210.

15. Kokmen E, Whisnant J, O’fallon W, Chu C-P, Beard C. Dementia after ischemic stroke: a population-based study in Rochester, Minnesota (1960-1984). Neurology. 1996;46(1):154-159.

16. Bornstein N, Gur A, Treves T, et al. Do silent brain infarctions predict the development of dementia after first ischemic stroke? Stroke. 1996;27(5):904–905.

17. Desmond DW, Moroney JT, Paik MC, et al. Frequency and clinical determinants of dementia after ischemic stroke. Neurology. 2000;54(5):1124–1131.

18. Zhou D. Study on frequency and predictors of dementia after ischemic stroke: the Chongqing Stroke Study. Annals of General Psychiatry. 2006;5(1):S195.

19. Sachdev P, Brodaty H, Valenzuela M, et al. Clinical determinants of dementia and mild cognitive impairment following ischaemic stroke: the Sydney Stroke Study. Dementia and geriatric cognitive disorders. 2006;21(5-6):275–283.

20. Gur A, Gücüyener D, Korczyn A, et al. Cerebral vasomotor reactivity and dementia after ischemic stroke. Acta Neurologica Scandinavica. 2010;122(6):383–388.

21. Allan LM, Rowan EN, Firbank MJ, et al. Long term incidence of dementia, predictors of mortality and pathological diagnosis in older stroke survivors. Brain. 2011;134(12):3716–3727.

22. Melkas S, Laurila JV, Vataja R, et al. Post-stroke delirium in relation to dementia and long-term mortality. International journal of geriatric psychiatry. 2012;27(4):401–408.

23. Brucki SMD, Machado MF, Rocha MSG. Vascular Cognitive Impairment (VCI) after non-embolic ischemic stroke during a 12-month follow-up in Brazil. Dementia & neuropsychologia. 2012;6(3):164–169.

24. Sibolt G, Curtze S, Melkas S, et al. Poststroke dementia is associated with recurrent ischaemic stroke. J Neurol Neurosurg Psychiatry. 2013;84(7):722–726.

25. Makin SD, Doubal FN, Quinn TJ, Bath PM, Dennis MS, Wardlaw JM. The effect of different combinations of vascular, dependency and cognitive endpoints on the sample size required to detect a treatment effect in trials of treatments to improve outcome after lacunar and non-lacunar ischaemic stroke. European stroke journal. 2018;3(1):66–73.

26. Kim TJ, Lee JS, Oh M-S, et al. Abstract WP552: Post-stroke Dementia Incidence and Risk Factors in Korea: Linked Big Data Between the Clinical Research Center for Stroke Data and the Health Insurance Review & Assessment Service Data Analysis. Stroke. 2019;50(Suppl_1):AWP552-AWP552.

27. Hachinski VC, Lassen NA, Marshall J. Multi-infarct dementia: a cause of mental deterioration in the elderly. The Lancet. 1974;304(7874):207-209.

28. Tatemichi TK, Desmond DW, Paik M, et al. Clinical determinants of dememtia related to stroke. Annals of Neurology: Official Journal of the American Neurological Association and the Child Neurology Society. 1993;33(6):568–575.

29. Snyder HM, Corriveau RA, Craft S, et al. Vascular contributions to cognitive impairment and dementia including Alzheimer’s disease. Alzheimer’s & Dementia. 2015;11(6):710–717.

30. Schneider J, Wilson R, Cochran E, et al. Relation of cerebral infarctions to dementia and cognitive function in older persons. Neurology. 2003;60(7):1082–1088.

31. Basso A, Capitani E, Luzzatti C, Spinnler H. Intelligence and left hemisphere disease. The role of aphasia, apraxia and size of lesion. Brain: a journal of neurology. 1981;104(Pt 4):721–734.

32. Mesulam MM. Large-scale neurocognitive networks and distributed processing for attention, language, and memory. Annals of Neurology: Official Journal of the American Neurological Association and the Child Neurology Society. 1990;28(5):597–613.

33. Gorelick PB, Brody J, Cohen D, et al. Risk factors for dementia associated with multiple cerebral infarcts: a case-control analysis in predominantly African-American hospital-based patients. Archives of neurology. 1993;50(7):714–720.

34. Inzitari D, Di Carlo A, Pracucci G, et al. Incidence and determinants of poststroke dementia as defined by an informant interview method in a hospital-based stroke registry. Stroke. 1998;29(10):2087–2093.

35. Thacker EL, McKnight B, Psaty BM, et al. Atrial fibrillation and cognitive decline: a longitudinal cohort study. Neurology. 2013;81(2):119–125.

36. O’Sullivan M. Leukoaraiosis. Practical neurology. 2008;8(1):26–38.

37. Caplan LR, Hennerici M. Impaired clearance of emboli (washout) is an important link between hypoperfusion, embolism, and ischemic stroke. Archives of neurology. 1998;55(11):1475–1482.

38. Ott A, Breteler MM, de Bruyne MC, van Harskamp F, Grobbee DE, Hofman A. Atrial fibrillation and dementia in a population-based study: the Rotterdam Study. Stroke. 1997;28(2):316–321.

39. Curb J, Rodriguez B, Abbott R, et al. Longitudinal association of vascular and Alzheimer’s dementias, diabetes, and glucose tolerance. Neurology. 1999;52(5):971–971.

